# Dynamics of COVID-19 under social distancing measures are driven by transmission network structure

**DOI:** 10.1101/2020.06.04.20121673

**Authors:** Anjalika Nande, Ben Adlam, Justin Sheen, Michael Z. Levy, Alison L. Hill

**Affiliations:** Program for Evolutionary Dynamics, Harvard University, Cambridge, MA, 02138; Department of Biostatistics, Epidemiology and Informatics, University of Pennsylvania, Philadelphia, PA 19104; Institute for Computational Medicine, Johns Hopkins University, Baltimore, MD 21218

## Abstract

In the absence of pharmaceutical interventions, social distancing is being used worldwide to curb the spread of COVID-19. The impact of these measures has been inconsistent, with some regions rapidly nearing disease elimination and others seeing delayed peaks or nearly flat epidemic curves. Here we build a stochastic epidemic model to examine the effects of COVID-19 clinical progression and transmission network structure on the outcomes of social distancing interventions. Our simulations show that long delays between the adoption of control measures and observed declines in cases, hospitalizations, and deaths occur in many scenarios. We find that the strength of within-household transmission is a critical determinant of success, governing the timing and size of the epidemic peak, the rate of decline, individual risks of infection, and the success of partial relaxation measures. The structure of residual external connections, driven by workforce participation and essential businesses, interacts to determine outcomes. We suggest limited conditions under which the formation of household “bubbles” can be safe. These findings can improve future predictions of the timescale and efficacy of interventions needed to control second waves of COVID-19 as well as other similar outbreaks, and highlight the need for better quantification and control of household transmission.

**Author Summary:** Social distancing is the main tool used to control COVID-19, and involves reducing contacts that could potentially transmit infection with strategies like school closures, work-from-home policies, mask-wearing, or lockdowns. These measures have been applied around the world, but in situations where they have suppressed infections, the effect has not been immediate or consistent. In this study we use a mathematical model to simulate the spread and control of COVID-19, tracking the different settings of person-to-person contact (e.g. household, school, workplace) and the different clinical stages an infected individual may pass through before recovery or death. We find that there are often long delays between when strong social distancing policies are adopted and when cases, hospitalizations, and deaths peak and begin to decline. Moreover, we find that the amount of transmission that happens within versus outside the household is critical to determining when social distancing can be effective and the delay until the epidemic peak. We show how the interaction between unmitigated households spread and residual external connections due to essential activities impacts individual risk and population infection levels. These results can be used to better predict the impact of future interventions to control COVID-19 or similar outbreaks

## Introduction

In less than five months the novel coronavirus SARS-CoV-2, the causative agent of COVID-19, has spread from an initial foci in Wuhan, China to nearly every corner of the globe. At the time of writing, over 1 million deaths had been reported, which will likely make this emerging virus the top infectious cause of death this year. Several clinical and epidemiological features of COVID-19 have contributed to its disastrous effects worldwide. The overlap in symptoms with many endemic and milder respiratory infections - such as influenza, parainfluenza, respiratory syncytial virus, and seasonal coronaviruses - make syndromic identification of cases difficult. The relatively high percentage of infected individuals who require hospitalization or critical care compared to seasonal respiratory infections has put an unprecedented burden on the healthcare systems of hard-hit regions. The important role of presymptomatic and asymptomatic individuals in transmitting infection makes symptom-based isolation less effective. Uncertainty about the case fatality risk from COVID-19 [1] and misguided comparisons to seasonal influenza contributed to sluggish responses in many regions, in contrast to previous outbreaks of SARS and MERS.

In the absence of either a vaccine or antiviral therapy, and given the continuing limitations in testing capacity in most regions, the main tools implemented worldwide to control the spread of COVID-19 have been “non-pharmaceutical interventions” including “social distancing”, isolation of cases, and quarantine of contacts. All of these measures are crude attempts to prevent the person-to-person contact that drives the transmission of respiratory infections, and have been used since antiquity in attempts to control outbreaks of plague, smallpox, influenza, and other infectious diseases [2,3]. Social distancing is a blanket term covering any measure that attempts to reduce contacts between individuals, without regards to their infection status. Within two weeks of identifying the original outbreak in Wuhan, a *cordon sanitaire* had been implemented around the entire Hubei province, prohibiting travel in or out of the region and requiring individuals to remain in their houses except to buy essential supplies. Elsewhere schools and universities have been closed, international travel has been limited, restaurants and retailers shuttered, mask-wearing encouraged or required, and stay-at-home orders put in place.

Mathematical models of COVID-19 transmission provided early support for the idea that social distancing measures could “flatten the curve” and reduce the potential for COVID-19 cases to overwhelm healthcare resources. An influential report from the Imperial College COVID-19 Modeling Team showed that suppression of the epidemic to levels low enough to avoid overflow of healthcare capacity would require an “intensive intervention package” that combined school closures, case isolation, and social distancing of the entire population, applied for the majority of time over two years [4]. Kissler *et al* also came to the conclusion that large sustained reductions in the basic reproductive ratio R_0_ (the average number of secondary infections generated by an infected individual) would be needed, even after accounting for the potential role of seasonality in transmission [5]. Many more forecasting models predicted dramatic decreases in the burden of COVID-19 if interventions were enacted (e.g. [6,7]). Real-time and retrospective analyses of the growth rate of cases and deaths have suggested that in some settings the epidemic eventually slowed after the implementation of strong social distancing measures (e.g. in Wuhan and other Chinese cities [8,9], in Hong Kong [10], across European countries [11], French regions [12], or some US states [13,14]).

The observed dynamics of COVID-19 outbreaks following social distancing policies have been inconsistent, unpredictable, and the source of much confusion and debate in the general public and among epidemiologists. Declines in cases and deaths have not occurred uniformly across regions and have often only occured after a long delay (**Figure 1**). The economic and social costs of these measures are immense: unemployment has surged, stock markets have plummeted, delivery of healthcare for non-COVID-19 conditions has been interrupted [15–19]. Social isolation also brings on or exacerbates mental health conditions. Weeks after implementing strong interventions, many regions have continued to see increases in daily diagnoses and deaths. Does this mean the interventions are not working? Since the political will to sustain strict social distancing measures is waning in many places, it is important to understand the expected timescale to judge success or failure. If stronger interventions - such as “shelter-in-place” orders or institutional isolation of mild cases - are needed to slow spread, when will we know this? What epidemiological and demographic features impact the timescale for epidemic waning, and how can we better predict the required duration of these measures for future outbreaks?

**Figure 1:**
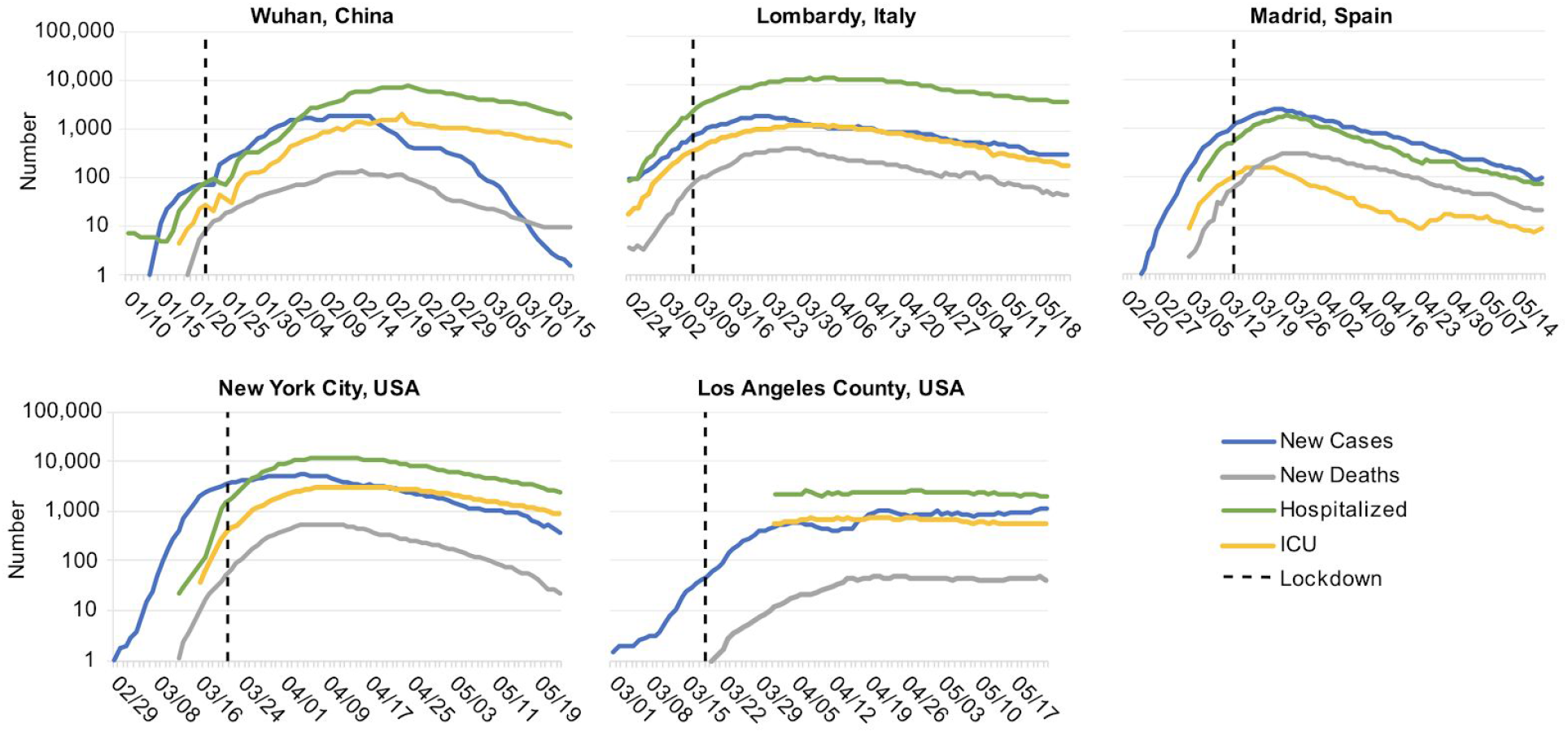
COVID-19 dynamics before and after lockdown interventions in five example regions. A) The city of Wuhan, China (8.5K km^2^, 11.1M ppl), B) The Lombardy region of Italy (23.8K km^2^, 10.1M ppl) C) The autonomous Community of Madrid in Spain (8.0K km^2^, 6.6M ppl) D) New York City in the state of New York, USA (1.2K km^2^, 8.2M ppl). E) The county of Los Angeles, California, USA (4.7K km^2^, 9.8M ppl). “New cases” and “New deaths” are daily numbers of new reports, averaged over a 7 day window centered on the current day. For Lombardy, New York, and Los Angeles, “Hospitalized” and “ICU” are the total number of patients currently in regular hospital care or critical care, respectively. In Wuhan, the same time series are the number of patients currently categorized as having “severe” or “critical” infection (using the same definitions as in our model). In Madrid, due to data availability, these series are instead the daily number of new admissions (with 7-day smoothing).

Social distancing measures reduce potentially-transmissive contacts occurring in schools, workplaces, social settings, or casual encounters, but they generally do so by confining individuals to their households without additional precautions. Thus, we would expect that the impact of social distancing measures might depend on the relative contribution of within-household transmission to disease spread, the distribution of household sizes, the number of households containing at least one infected individual at the time an isolation measure is enacted, and the amount of residual contact between households for the duration of the intervention. What do we know about these factors for COVID-19 or respiratory infections more generally, and how do they interact to determine epidemic dynamics after an intervention?

In this paper we examine the impact of COVID-19 clinical features and transmission network structure on the timing of the epidemic peak and subsequent dynamics under social distancing interventions. Using data from large-scale cohort studies, we parameterize a model tracking the progression of COVID-19 infection through different clinical stages. We combine this with data-driven transmission networks that explicitly consider household vs external contacts and how they are differentially altered by social distancing measures. We consider various scenarios for the efficacy of interventions in reducing contacts, heterogeneities in their adoption in different demographic groups, the relative role of transmission in different settings, and the timing of partial or complete relaxation of isolation measures. We evaluate both population-level outcomes as well as determinants of individual risk of infection. Our results show that even following the implementation of strong social distancing measures, the epidemic peak can occur weeks to months later, and the decline in cases can be extremely slow. The efficacy of within-household transmission plays a critical role in the timescale and overall impact of these measures. These findings provide an impetus for continued adherence to social distancing measures in the absence of immediate results, can inform planning for hospital capacity, and suggest that retrospective efforts to assess the efficacy of different intervention policies should account for these expected delays.

## Methods

### Modeling the spread and clinical progression of COVID-19

We modified the classic SEIR compartmental epidemiological model to describe the dynamics of COVID-19 infection (**Supplementary Methods, Figure 2A**). After infection, individuals pass through an ∼ 5 day incubation period before developing asymptomatic or mild infection, which could include fever and cough or other symptoms. This stage lasts ∼ 1 week and individuals are infectious for this duration. A portion of individuals progress to “severe infection”, which is typically characterized by pneumonia requiring hospitalization, and we assume averages 6 days. Some individuals progress further to “critical infection”, which requires ICU-level care that often includes mechanical ventilation, and some of these individuals eventually die (after ∼ 8 days of critical care), leading to an ∼2% case fatality risk. At each stage, individuals who don’t progress or die, recover and are assumed to be immune for the duration of the outbreak. The duration of each stage of infection is assumed to be gamma-distributed with mean and variance taken from the literature. Infectious individuals can transmit to any susceptible individuals with whom they are in contact, with a constant rate per time for the duration of their infection. With our baseline parameters, the doubling time of infection is ∼ 4 days, the basic reproductive ratio R_0_ is ∼3, and the serial interval is ∼8 days, in agreement with epidemiological studies of COVID-19. A detailed description of the clinical definitions of different infection stages, the model behavior, and the model parameters and references are given in the Methods.

**Figure 2:**
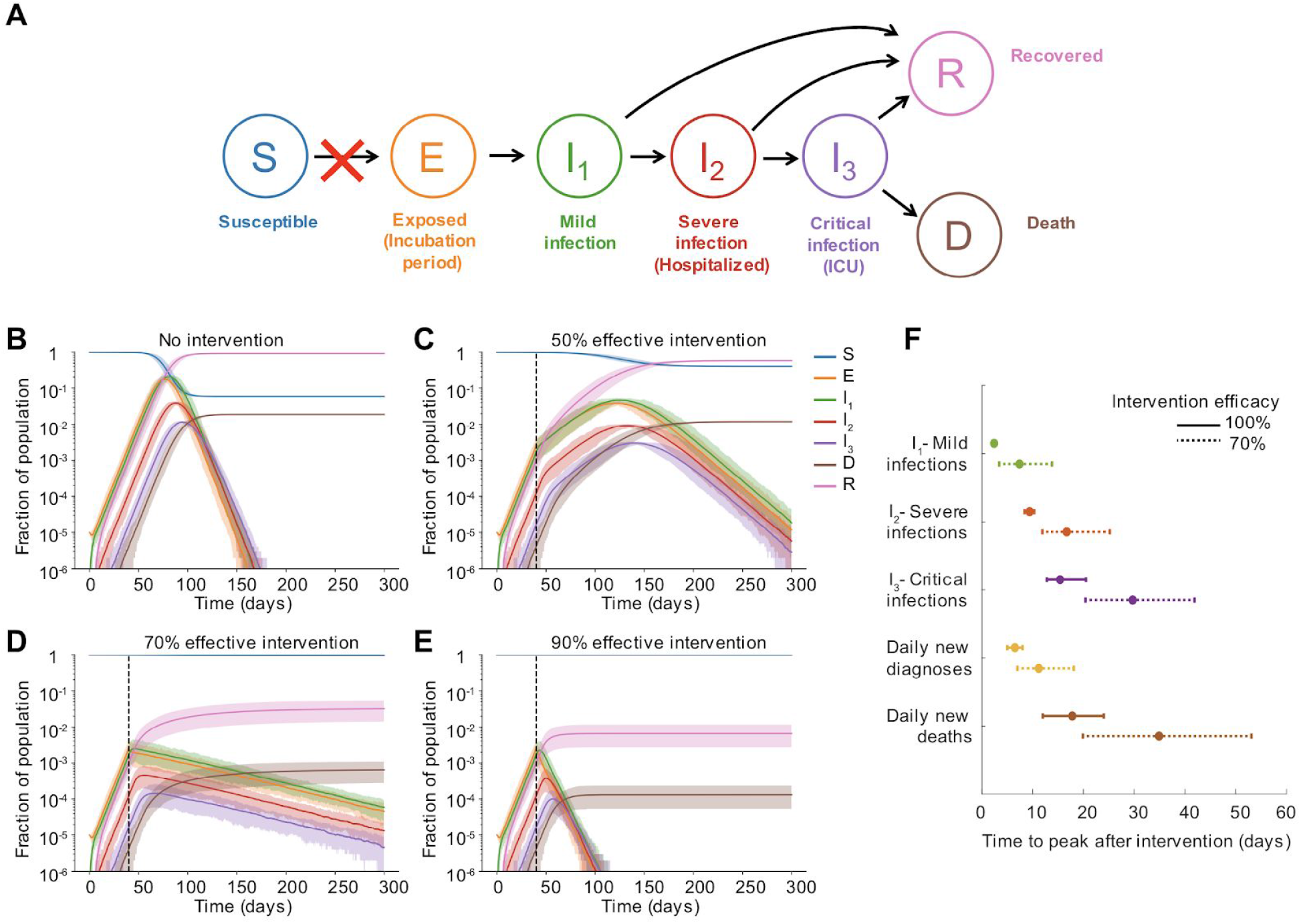
Dynamics pre and post social distancing intervention in well-mixed populations. A) Model of COVID-19 clinical progression and transmission. The model is described in the text and detailed in the Methods. Social distancing interventions (red X) reduce the rate of transmission and the generation of new infections. B-E) Simulated time course of the population level prevalence of each clinical stage of infection under different intervention efficacies. The intervention was implemented on day 40. Solid line is mean and shaded areas are 5th and 95th percentile. Black dotted line shows the time the intervention began. F) Time to peak of different infection stages, measured as days post-intervention. The first three quantities are peak prevalence levels (I_1_, I_2_, I_3_), while the latter two are peak daily incidence values. We assume that cases are diagnosed only at the time of hospitalization. Daily incidence values were first smoothed using moving averages over a 7 day window centered on the date of interest. Bars represent 5th and 95th percentile.

We then simulate infection spreading stochastically through a fixed, weighted contact network with one million nodes. The population size is chosen to represent a typical metropolitan area. As a baseline scenario, we consider a simple approximately well-mixed population where anyone can potentially transmit the virus to anyone else in the population. To more accurately capture human contact patterns, and how they are altered by social distancing measures, we constructed multi-layer networks describing connections within households and external connections (**Supplementary Methods, Figure 3A**). Each individual was assigned to a household and connected to everyone in their house. Household size distributions were taken from the 2010 United States census (average household size *n*_*HH*_∼2.5, full distribution shown in **Figure 3B**). External connections were constructed by connecting individuals to people in other households. The distribution of the number of external connections was taken from detailed contact surveys that recorded daily interactions amenable to transmission of respiratory infections (average *n*_*EX*_∼7.5, standard deviation 2.5) [20,21]. As a baseline case we constructed “two-layer” networks assuming these external connections were random, whereas later in the paper we consider more complex and realistic “five-layer” network structures. While these data sources inform the *number* of contacts, the probability of infection depends both on the number of unique contacts and on the time spent together and the intensity of the contact, which can be represented by weights in the network. We hypothesized that household and external contacts could have different effective weights. For example, individuals may spend 8-10 hours a day with coworkers or classmates, but only a few waking hours with household members, and so external contact could have higher weights. Alternatively, individuals may have more intense physical contact with household members, such as children or spouses with whom co-sleeping can occur. Since these weights are unknown, we considered a range of scenarios for the relative weights of household (*w*_*HH*_) and external (*w*_*EX*_) contacts, keeping the total transmission intensity (basic reproductive ratio *R*_*0*_) constant. These scenarios result in different observed values of the household “secondary attack rate” (the probability a single index individual infects any given household contact) (**Figure 3K**). We also hypothesized that when individuals are isolated in their homes as a result of social distancing measures (e.g. school closures or work-from-home mandates), they may be spending significantly more time with household members and thus have a higher transmission rate. We modeled this by allowing the weight of household contacts to increase during an intervention.

**Figure 3.**
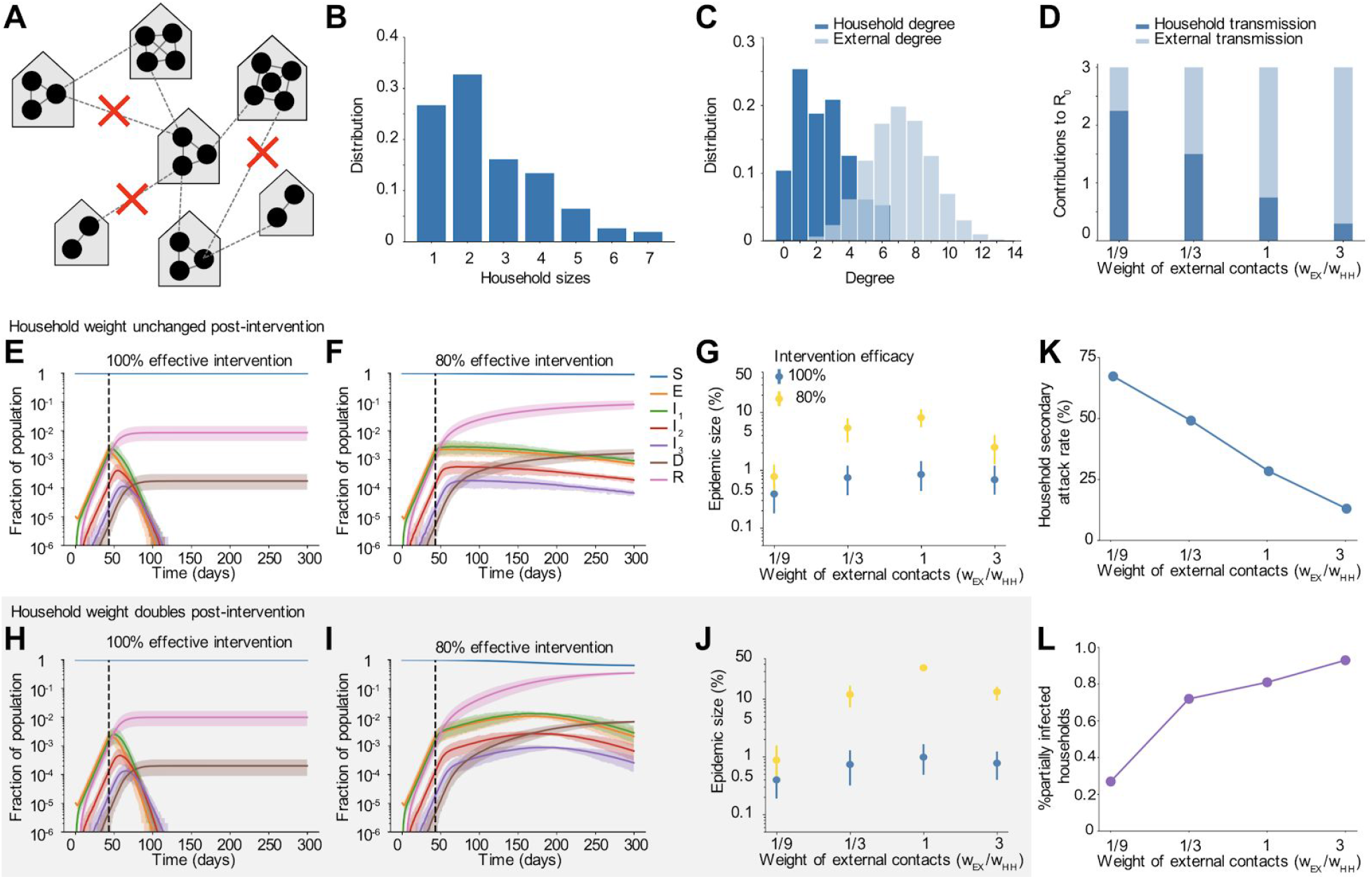
Dynamics pre and post social distancing interventions in network-structured populations with household and external transmission. A) Multi-layer network of transmission. Individuals have contacts within their households and with others outside the household. Household and external contacts may have different weights (e.g. different likelihood of transmission), due to for example different levels of physical contact or time spent together per day. Social distancing interventions (red X) remove or decrease the weight of external contacts. B) Distribution of household sizes. C) Distribution of the # of contacts (degree) within the household and outside the household. D) The contribution of household and external spread to the total R_0_ value as a function of the relative weight of external contacts. E)-F) Simulated time course of different clinical stages of infection under an intervention with efficacy of 100% (E) or 80% (F) at reducing external contacts, when household and external contacts have equal weight. Black dotted line shows the time the intervention began. G) The role of the relative importance of household vs external contacts in determining the outcome of the intervention, measured by the size of the epidemic. Epidemic final size is defined as the percent of the population who have recovered by day 300. H-J) Same as above but under the scenario where the weight of household contacts doubles post-intervention (*w*_*HH*_ → 2*w*_*HH*_, due to increased time spent in house). K) The household secondary attack rate, defined as the probability of transmission per susceptible household member when there is a single infected individual in the house, as a function of the relative weight of external contacts. L) The percent of households which are “seeded” with infection at the time the intervention was implemented (i.e. have at least one infected individual). In all scenarios the overall infection prevalence at the time intervention was started was identical.

We model the implementation of social distancing measures by reducing the weight of all external contacts (or all contacts in the well-mixed model) by a fixed % that we term the “intervention efficacy”. Alternatively, we could randomly remove a fixed % of contacts, but the results are very similar (see **Methods**). Our model is similar to other models that have been used to describe the spread of COVID-19. A unique feature of our model is that it simultaneously captures the clinical progression of COVID-19 (as opposed to simpler SEIR models), a reasonable approximation of contact network structure (as opposed to well-mixed models), and realistic distributions of the durations of states (as opposed to continuous-transition models which assume exponentially-distributed durations, and lead to unrealistically long tails in infection after strong interventions). We can simulate infections for the duration of the epidemic in less than 1 minute on a single GPU, in populations of a million.

## Results

### Observed COVID-19 dynamics following social distancing interventions

To characterize the dynamics of COVID-19 following social distancing measures, we chose five regions from around the world with large outbreaks: the city of Wuhan, China, the Lombardy region of Italy, the Community of Madrid in Spain, New York City in the state of New York, USA, and the county of Los Angeles, California, USA (**Figure 1**). These regions each implemented strong “lockdown” measures (aka “stay-at-home” or “shelter-in-place” orders) within 3 weeks of their first reported COVID-19 case and provided data not just on cases and deaths but also on cases requiring hospitalization and ICU-level care (see **Supplementary Methods**). In each setting, there was a long delay between the implementation of social distancing and the peak incidence of cases (1.5-3 weeks) and deaths (2-3 weeks), or peak occupancy in hospitals and ICUs (∼1 month). The timescale of the eventual decline in cases post-peak was much slower than the initial increase in cases in all regions, with a half-life between 10 and 24 days in all regions except Los Angeles, where the outbreak appears to have approximately plateaued but not yet begun decreasing. The goal of this paper was to understand whether the clinical progression of COVID-19 and transmission network structure could explain these types of post-intervention dynamics.

### Prolonged clinical progression of COVID-19 leads to delay until decline in cases and deaths following an intervention

We first considered the role of the clinical features of COVID-19 alone, in the delay from implementation to peak infections and deaths, by simulating our model in an unstructured population. The intervention was implemented when cumulative reported cases were ∼200 per million and deaths ∼5 per million (total infected ∼1%), mirroring the timing of stay-at-home orders across major US metropolitan areas (see **Supplementary Methods**). While we expect the number of new infections to begin decreasing immediately, newly infected individuals in the “exposed class (E)” (incubation period) cannot generally be tracked, since they are asymptomatic and not yet shedding enough virus to test positive. Instead, later stages of infection are monitored.

We found that under a perfect intervention, we expect ∼2 days delay until the peak prevalence of mild infections, ∼9 days for severe infections, and ∼15 days for critical infections, suggesting that the requirements for healthcare capacity may peak quite a bit after implementation (**Figure 2**). In the more realistic scenario where the intervention is imperfect (70% effective), these timelines are significantly extended, for example to ∼7, 17, and 30 days for mild, severe, and critical infections respectively. In most regions, individuals are reported at the time of diagnosis, and not tracked until recovery, and so case counts can only be used to track *incidence* rates, not prevalence levels. We consider a region where infections are only counted upon hospitalization (progression to severe class), and then find that peak incidence of cases occurs 7 and 11 days after an intervention that is 100% or 70% effective. Daily deaths peak much later : after 18 days (100% effective) to 35 days (70% effective). Under our parameter values, a 50% intervention “flattens the curve” but does not prevent spread, and incidence cases and deaths don’t peak until 13 and 15 weeks after the intervention, respectively. The total percent of the population infected over the course of the whole epidemic was reduced from ∼92% to ∼0.6% with a 100% effective intervention, but only to 58%, 3%, or 0.65% with a 50, 70% or 90% effective intervention.

The exact timings that we report here depend on the assumptions of our model, in particular, the average duration of each stage of infection (see **Supplementary Methods** for details) as well as on the epidemic growth rate pre-intervention (it takes longer for epidemics that were growing faster to peak and begin declining). However, the qualitative finding that peaks in case counts, hospitalizations, and deaths can be significantly delayed beyond when an intervention is implemented is a general finding for models tracking the natural history of COVID-19. Note that in our model, we assume that the intervention is adopted the same day it is instituted, whereas in reality, there may be a further delay until individuals are able to comply with the intervention.

### The relative contribution of household and external spread influences outcome of interventions

We hypothesized that the continual spread of COVID-19 within households after the implementation of social distancing measures could further delay peak cases and deaths, and increase the number of people infected despite the intervention. Using our network-structured model (see **Methods**) for household and external contacts, we simulated the implementation of interventions of increasing efficacy under different assumptions about the relative weight of the household vs external contacts. In addition, we examined the impact that the increased time spent with household members (and hence an increased transmission potential) after stay-at-home policies begin could have on the outcome of an intervention and the timescale for disease elimination (**Figure 3**).

With our baseline assumption that household and external contacts had equal weight, we observed that cases declined rapidly under very strong interventions (**Figures 3E and H**), while imperfect interventions (e.g. ∼80%) often resulted in very gradual decreases in cases over many months (**Figure 3F**). In both scenarios the eventual fraction of the population infected was dramatically reduced compared to the no intervention case, but these long timescales likely mean that costly social distancing policies cannot be maintained long enough for suppression of the epidemic to occur. This slow decline could be further compromised if the risk of transmission within a household increases under stay-at-home policies (**Figure 3I**). In this case the epidemic could continue to increase for months post-intervention before eventually declining, albeit still to a much lower final size than in the absence of interventions.

When the outcome of an intervention was measured by the total fraction of the population infected over the course of the outbreak, we found that there was a surprisingly complex relationship between the relative contribution of household and external contacts to transmission, and the intervention success (**Figures 3G and J**). Keeping the total R_0_ constant, social distancing interventions are most effective when either external contacts have very high weights or when they have very low weights. In the former case (high external weight + low household weight), most of the pre-intervention transmission comes from outside the household, and the intervention is very effective at blocking this transmission (**Figure 3D**,**K**). At the time the intervention is implemented, many households are “seeded” with infections that originated outside the house (**Figure 3L**), but after the intervention, household transmission alone is not effective enough to lead to a new generation of infections in most houses, without seeding from the outside (i.e intervention efficacy <100%). When external contacts have low weight, the intervention is highly effective but for a different reason. Most transmission is inside the household and can continue post-intervention (**Figure 3D**,**K**), but very few households are seeded with infections (**Figure 3L**). The weak inter-household contacts are further weakened by the intervention and spillover between households is unlikely, meaning that the infection quickly burns through susceptibles within a household then dies out.

In the intermediate regime, where household and external contacts have approximately equal weight, social distancing interventions are less effective, and are very sensitive to imperfect efficacy. For example, when external contacts have ∼1/3 the weight of household ones, each type of contact contributes equally to the overall pre-intervention *R*_*0*_ (since there are ∼3x the number of external contacts as household ones). With a 100% effective intervention, the final epidemic size is ∼0.7%, but rises to ∼7% with a 80% effective intervention (**Figure 3G**). The combination of enough household spread (*R*_*0*_ ^*HH*^ *>1*) to allow efficient transmission post-intervention within “seeded” households and enough external spread (*R*_*0*_ ^*EX*^ *>1*) to seed households before the intervention is implemented to allow post-intervention spillover of infections to other households is the most difficult case for control. These effects are exacerbated if we assume household transmission rates (contact weights) can increase post-intervention (**Figure 3J**). For an 80% effective intervention, the final epidemic size can be 5-10 - fold higher than expected due to increased chance of within-household transmission. We repeated these simulations with a hierarchically-clustered external layer (see **Methods**) to check the robustness of the trends to details in the large-scale clustering of the transmission network (**Figure S1, S2**). We found the trends to be preserved, with the most noticeable difference being in the number of houses “seeded” with infection at the time of intervention.

### Residual household transmission can further delay time to see the impact of an intervention

We found that the expected time to peak infections and deaths after a social distancing intervention was implemented could be increased dramatically when we accounted for household structure, and was sensitive to the relative importance of household and external contacts before and after the intervention (**Figure 4**). Under a 100% effective intervention (**Figure 4A**), the delays to peaks were driven mainly by the clinical progression alone, similar to the case of the well-mixed population, but were slightly extended due to residual spread restricted to a single household. In simulations it took around 2 weeks until peak hospitalizations and 3 weeks to peak critical care cases or daily deaths. However, under an imperfect but still strong intervention (e.g. 80% effective), the times to peak were much longer and sensitive to the relative weights of the external and household contacts (**Figure 4B**). Delay to peak cases was longest in the intermediate regime where external and household contribution to transmission was approximately equal. For example, when external and household weights were equal, it took an average of ∼ 5.5 weeks to reach peak cases with mild symptoms, ∼ 7 weeks until peak cases hospitalized with severe infection, and ∼ 8.5 weeks to the peak of cases in critical care. The daily incidence of new deaths didn’t peak for ∼ 10 weeks.

**Figure 4:**
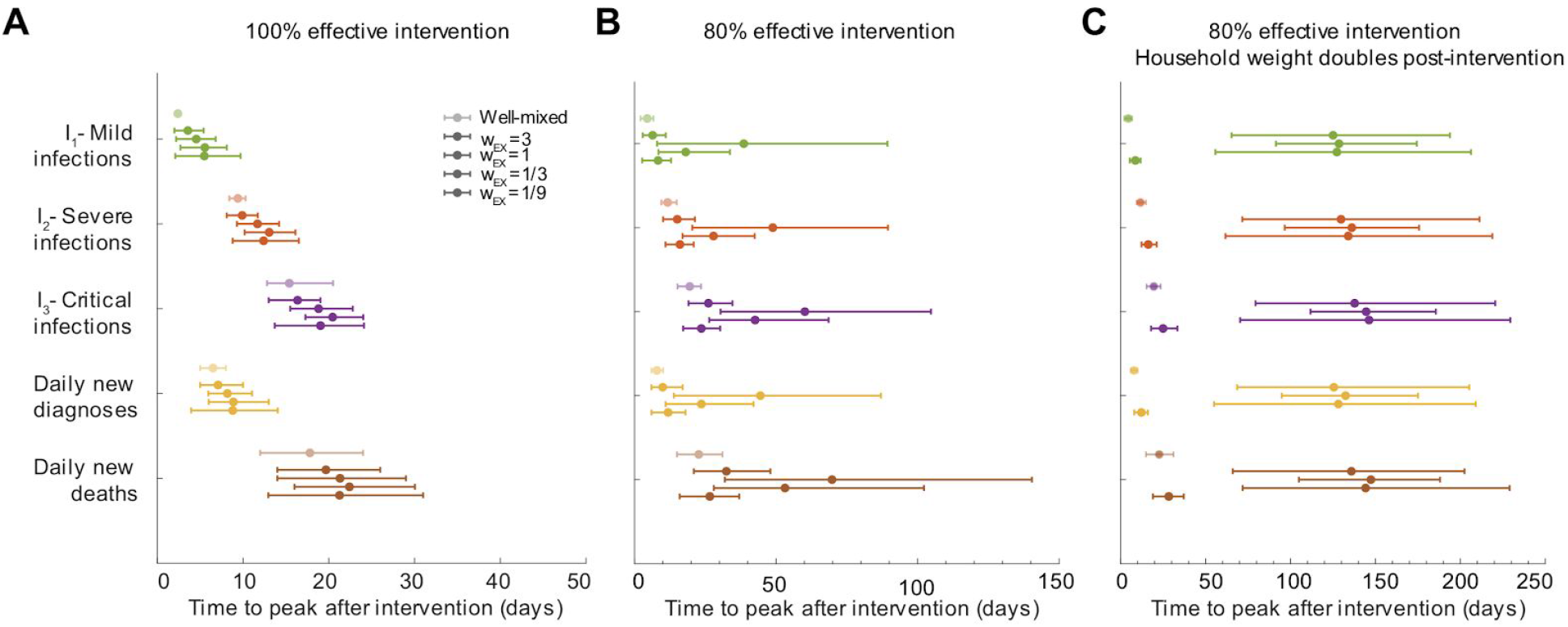
Time to epidemic peak after social distancing interventions depends on the relative roles of household and external transmission. A-C) Time to peak of different infection stages, measured as days post-intervention. A) Social distancing intervention with 100% efficacy at reducing external contacts (or all contacts in the case of a well-mixed network). B) Social distancing intervention with 80% efficacy. C) Social distancing intervention with 80% efficacy, and assuming that household weights double post-intervention (*w*_*HH*_ → 2*w*_*HH*_, due to increased time spent in the home). The first three quantities are peak prevalence levels (I_1_, I_2_, I_3_), while the latter two are daily incidence values. We assume that cases are diagnosed only at the time of hospitalization. Daily incidence values were first smoothed using moving averages over a 7 day window centered on the date of interest. Bars represent 5th and 95th percentile. For each clinical stage included (each different color), the lighter-colored data point is the comparison to the well-mixed population, then the other points are for decreasing contributions of external connections and increasing role of household transmission.

The delays in time to peak were less extreme if external contacts had very high or very low weights relative to the weight of household contacts (**Figure 4B**). In the case of very high external weight, most individuals were infected from contacts outside their household before the intervention (**Figure 3L**). Household spread is relatively inefficient, and has only a minor contribution to the baseline *R*_*0*_ value (**Figure 3D**). In most households, there is no further spread after the intervention is implemented. As a result, the epidemic peaked sooner: peak daily deaths occured an average of ∼ 3 weeks post 100% effective intervention and ∼ 5 weeks post 80% effective intervention. On the other hand, when external weight is a lot lower as compared to the household, only a small fraction of households are seeded with infection by the time intervention is started (**Figure 3L**). Intervention is very effective at suppressing external transmission and so, even though household transmission continues during intervention it can not spill over between households. This causes the epidemic to peak sooner as susceptibles in households get infected quickly and then the infection dies out. On average, peak daily deaths occurred ∼ 3 weeks (100% effective intervention) and ∼ 4 weeks (80% effective intervention) post intervention.

These results were exacerbated if we assumed that the importance of household contacts increased post-intervention (**Figure 4C**), due to increased time spent in close quarters. In that case, peaks increased to up to ∼ 6 months for cases in critical care and daily deaths under an 80% intervention. With higher household weights, the efficacy of spread within a household was stronger, making new generations of infection post-intervention very likely to occur in households with at least one case. Then, these household infections are more likely to spill over into other households, even when most external contacts are eliminated by the intervention. Together, these effects allow for multiple generations of transmission to persist even after a strong intervention.

In many regions around the world, the effect of social distancing interventions is monitored in real-time using estimates of the time-dependent reproduction number *R*_*t*_ (e.g. [22]). We applied standard procedures for calculating *R*_*t*_ [23] to the incidence data from our simulations, and using the time at which *R*_*t*_ first crossed the threshold of 1 as a measure of the delay, we found that the trends agreed with those reported for the epidemic peak (**Figure S8**).

### Clustered adoption of social distancing measures can further compromise efficacy

Our results so far have assumed that external contacts in the transmission network are random connections between pairs of individuals in the population, and that a social distancing intervention results in a uniform random reduction or deletion of these connections. In reality, human contact networks tend to be highly structured, with groups of individuals with high levels of interconnectedness and large variation between individuals in total contacts (e.g. [24,25]). Moreover, we don’t necessarily expect adherence to social distancing measures to be random. For certain occupations or in certain demographic groups, individuals are less likely to be able to work-from-home or otherwise reduce contacts outside the home. This can lead to clusters of individuals among whom contacts remain high despite interventions. We hypothesized that this clustered adoption of social distancing measures could lead to more residual transmission, longer times to peak cases and deaths, and longer times to eradicate infection from a given region.

To examine these effects, we constructed more realistically-structured, age-segregated external contact networks. The population was divided into four broad age groups: preschool-aged, school-aged, working-aged and elderly. Based on large-scale contact surveys and other modeling studies [20,21,25–27], we broke down external contacts into four different layers - school, work, social and community (**Figure 5A**). Age groups determined network membership. School and work layers consisted of connections between individuals only belonging to the school-aged and working-aged groups respectively. Individuals belonging to all age groups were part of the social and community layers. We used a variety of data sources to construct the networks for each layer with degree distributions (both mean and variance in # of contacts) as well as levels of clustering (aka transitivity, a measure of interconnectedness) that matched data (see Methods). We assumed that during a social distancing measure, school contacts were completely removed, and that work, social, and other contacts were reduced by an amount equal to the intervention efficacy. For work contacts, we also considered the case where edges weren’t removed at random, but instead, certain “workplaces” were completely dissolved, whereas others remained (**Figure 5C**, top). With this implementation, the levels of clustering in the external network was high both before and after an intervention. Other studies have shown that such clustered adoption of preventive behavior can lead to lower than expected efficacy of vaccines and mass drug administration [28–31].

**Figure 5:**
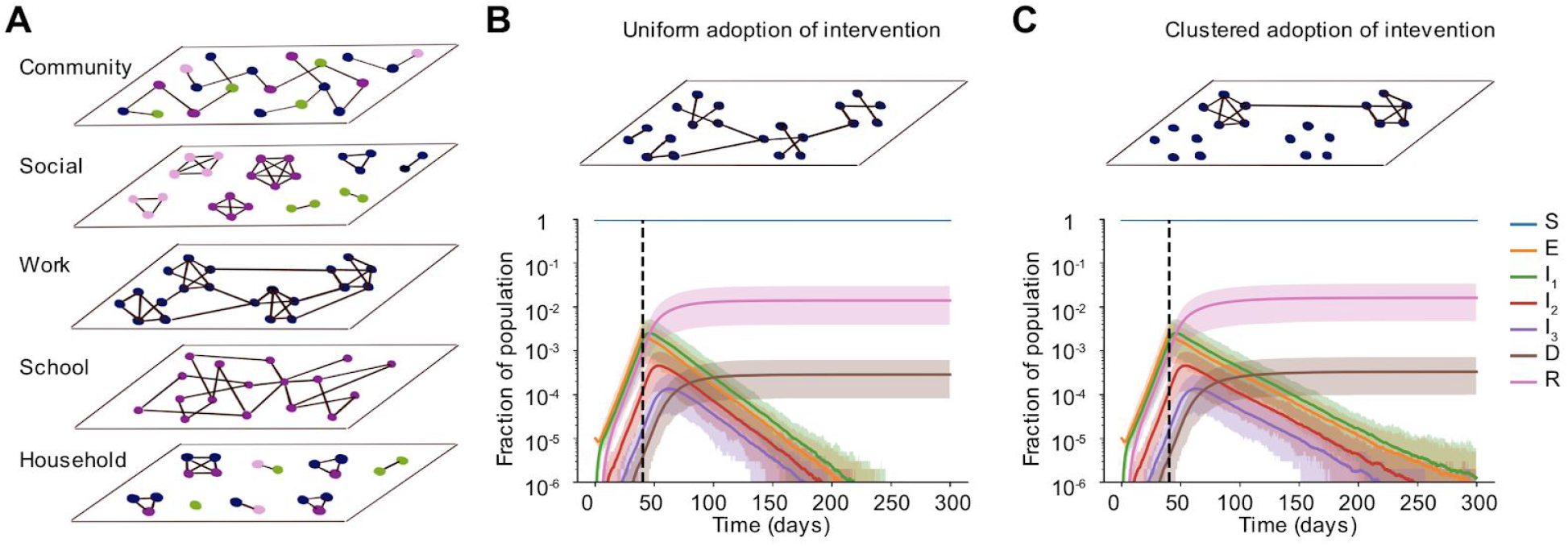
Clustered vs uniform adoption of social distancing measures. A) Schematic of the multi-layer network created to more realistically capture non-household contacts and how they are altered by social distancing measures. In each layer, the degree distribution and level of clustering were chosen to match data. The “community” layer represents any other contact not fitting in the other four categories. Colors of nodes represent four broad age groups that determine network membership and structure: preschool-aged (pink), school-aged (purple), working-aged (blue) and elderly (green). B) - C) Simulated time courses of infection in the presence of social distancing intervention with random (B) vs clustered (C) adherence to measures. In both cases, all school connections were deleted post-intervention and 85% of connections were uniformly deleted at random in the social and community layers. In B) 85% of work connections were uniformly deleted whereas, in C) 85% of workplaces were dissolved, leading to clusters of disconnected vs connected individuals in the work layer of the network. The effective intervention efficacy for all layers combined was ∼ 88% in both scenarios. Black dotted line shows the time the intervention began.

We found that when the intervention efficacy was high, most outcomes were surprisingly not worse under this clustered adoption (**Figure 5**). Time to peak cases, hospitalizations and deaths were similar under random deletion of edges (**Figure 5B**, bottom) and under the correlated deletion scheme (**Figure 5C**, bottom). However, we found that the time until infection was eliminated from the population was much longer: increasing from ∼ 180 days to ∼ 220 days for population sizes of a million. For a less effective intervention, the difference in outcomes for the two deletion schemes was more prominent (**Figure S3**). Under clustered adoption, the epidemic plateaued and took much longer to decline compared to the case of uniform adoption where decline began immediately. We again performed a sensitivity analysis to check the robustness of these findings to meso-scale clustering of contacts in the network (see Methods) and found the trends very similar (**Figure S4**). These findings suggest that targeting demographic groups like essential workers, where pockets of infection might persist, with more aggressive cases-based measures and contact tracing may be necessary to reach elimination goals faster.

### Individual risk of infection depends on household size and occupation

So far our evaluations of social distancing measures have focused on population-level outcomes such as the timing of the epidemic peak and the overall fraction of the population infected. However, these findings mask significant heterogeneity in individual risk. From our simulations, we extracted the individual probability of infection as a function of household size (**Figure 6A**), as well as in relation to the external contacts maintained after an intervention (**Figure 6B**). We found that the risk of infection increased dramatically with the household size: with our baseline parameters, it ranged from <0.2% for individuals living alone to 5.4% for households of size 7 (**Figure 6A**). These differences occurred independently of the relative weight of household vs external contacts. The supra-linear increase in risk with household size is driven by the fact that in larger households there is both more risk of seeding of infection from outside, as well as more individuals to spread to within the household leading to less chance of extinction of spread.

**Figure 6:**
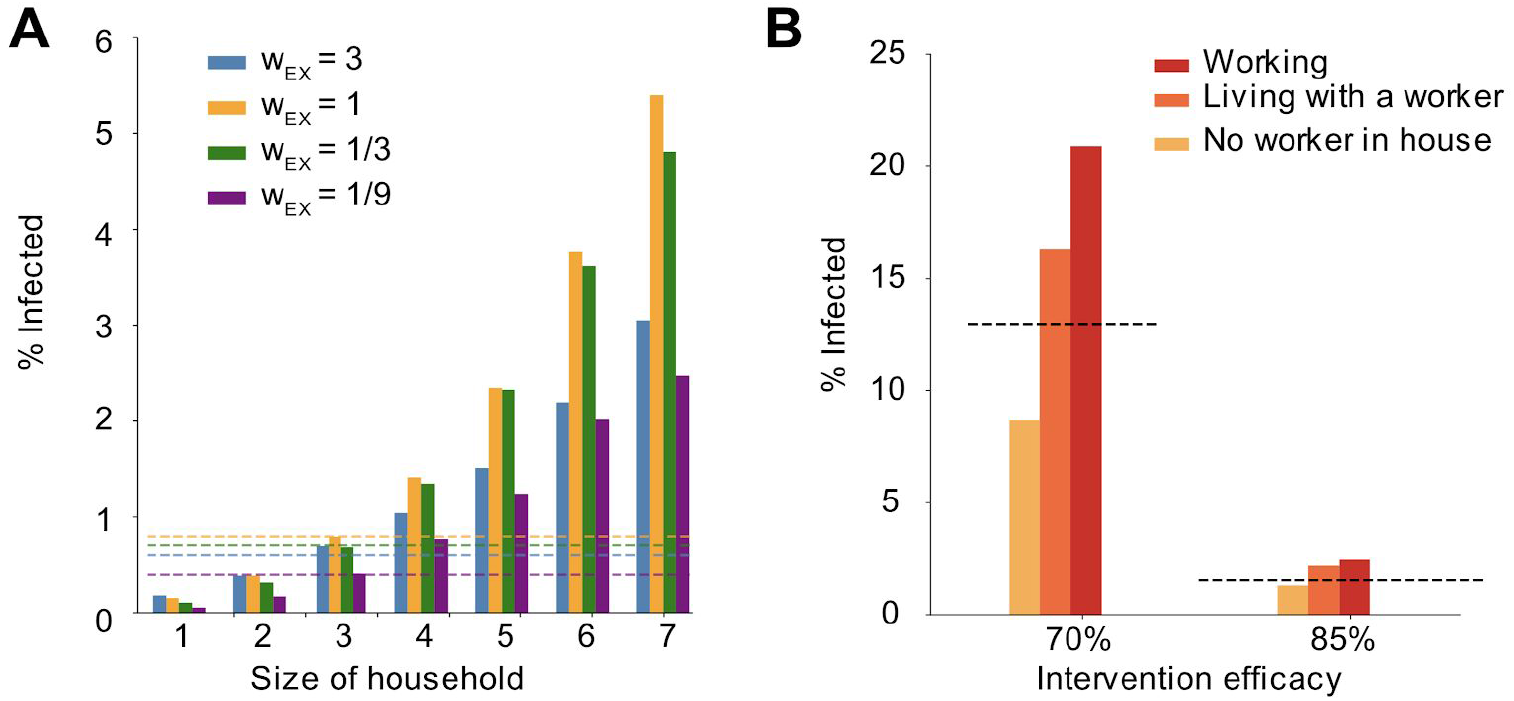
Individual risk of infection depends on household size and worker status. A) Risk of infection versus household size in simulations. Risk of infection was calculated after 300 days, with 100% intervention efficacy. Bar colors represent different relative weights of external contacts (compared to household contacts). Dotted lines are the population level average infection levels for the same scenarios. B) Risk of infection versus worker status. A “worker” is defined as someone with an occupation in which they continue to work outside the home despite social distancing measures. Categories include being a worker yourself (red), living in a household with at least one other individual who is working (orange), or having no workers in the house (yellow). As a comparison the population average risk is shown (dotted line). Interventions that reduce the overall number of people working outside the home by 70% and 85% are shown (in all cases all schools are assumed to be closed and the same percent of social and community contacts are removed).

We also examined the increased risk faced by “essential workers”, or others who maintained contacts in their “work” networks during the time social distancing measures were in place (**Figure 6B**). Under more extreme distancing (∼85% reduction in contacts), the relative risk of infection among workers relative to the population average was 1.6, while for individuals not working themselves but living in the same household as someone who was working was 1.4. In comparison, individuals belonging to households with no workers had a relative risk of 0.8. For a less effective intervention (∼70%), these values were 1.6, 1.3 and 0.7 respectively. These findings highlight the risk faced by communities in which larger households are common and/or in which more individuals per household may maintain external connections despite social distancing measures.

### Expanding circles can be safe partial relaxation strategies only under certain conditions

As a step towards relaxing social distancing measures in settings where the incidence of cases and deaths has stabilized or is declining, some regions are proposing partial relaxation strategies whereby groups of households merge to form larger “expanded circles” or “bubbles”, but still minimize external contacts [32,33]. Such multi-household groups could have enormous social benefits, such as providing childcare relief and improving productivity of working parents, and reducing the mental health toll of social isolation. To examine when this strategy could be safely implemented without risking a rebound in cases, we randomly joined households 1, 2, or 3 months after the implementation of a strong social distancing measure (80 or 90% effective) (**Figure 7**).

**Figure 7:**
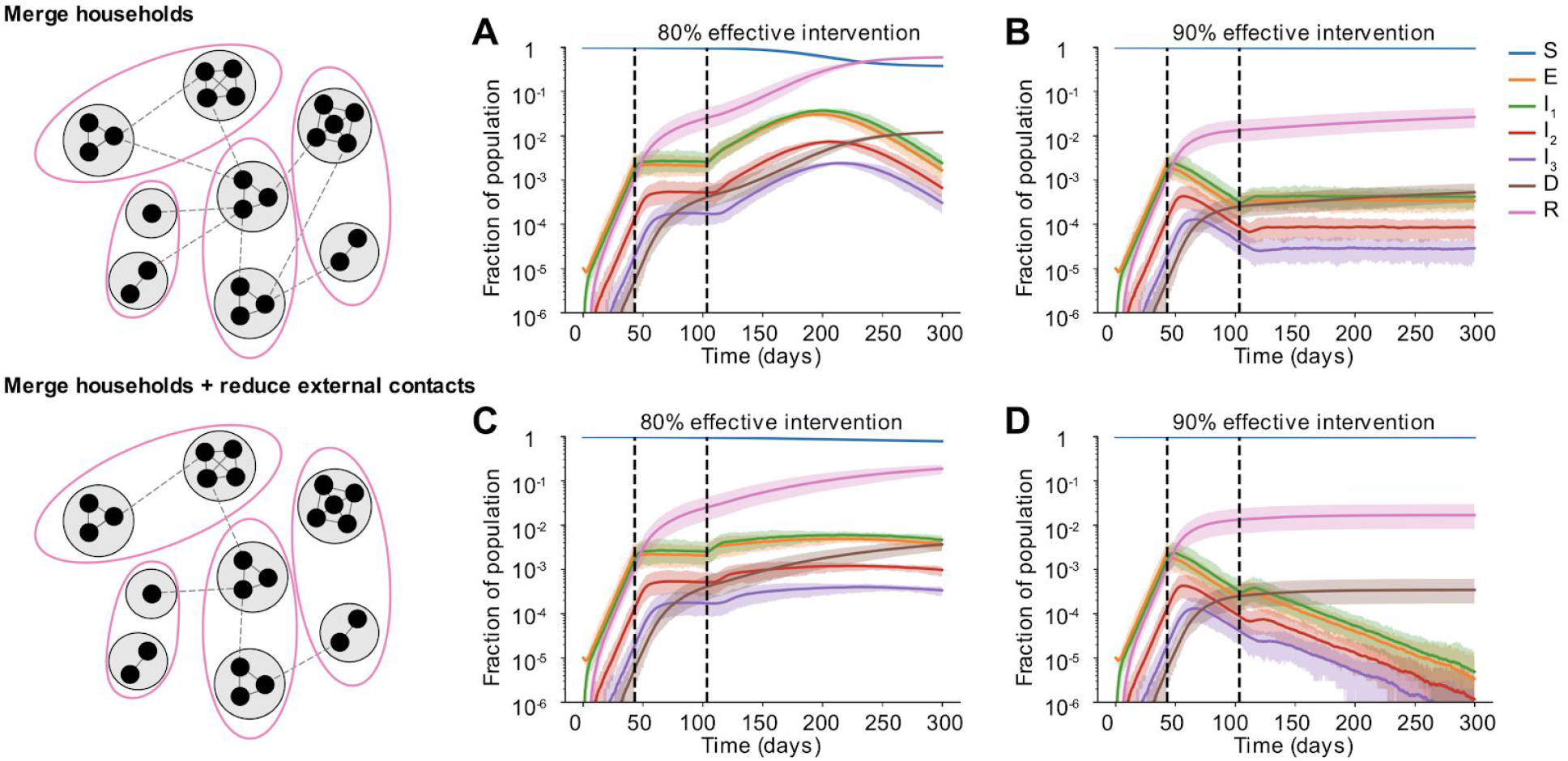
Effect of partially relaxing intervention by forming household bubbles. Some time after a social distancing intervention was implemented, each household merges with another random household. In each resulting two-household “bubble”, all individuals are connected to all other individuals. A)-D) Simulated time courses of infection before and after social distancing interventions (with 80% vs 90% intervention efficacy) and after partial-relaxation by household merging. Top row: External contacts of individuals were unchanged after two households were merged, such that overall number of contacts increased. Bottom row: External contacts for individuals were reduced after two households were merged, such that overall number of contacts remained unchanged. In all cases, intervention was started 43 days after the onset of the epidemic (first black dotted line) and was relaxed after two months (60 days, second black dotted line).

We found that these household-merging strategies could be safe only if a few criteria were met. Firstly, they must be applied in the context of steadily declining cases and deaths (**Figures 7B and D**). In situations where infection levels had stabilized but were barely declining, forming bubbles always led to at least some resurgence of cases which returned to or exceeded peak levels (**Figures 7A and C**). Secondly, household bubble formation should ideally be accompanied by a further decrease in contacts outside the house (for example, only one grocery trip per dual-family household instead of two) and a redistribution of the effective number of household contacts instead of allowing them to double (for example, by spending time with subsets of the dual household instead of all time as a complete group). Otherwise, a previously declining epidemic could instead stabilize at a persistent level (**Figure 7B**), or an otherwise stable epidemic could temporarily resurge (**Figure 7A**). When resurgence occurred it took 1-4 weeks to see noticeable increases in hospitalizations or deaths. We did not find a strong dependence on the timing of household bubble formation. As before, we also tested the robustness of these results to details in the large-scale clustering of the network by using a metapopulation model that incorporates the notion of “neighborhoods” (see Methods). We found that the trends were unaffected independent of whether the merged households belonged to the same (**Figure S6**) or different neighborhoods (**Figure S5**).

Clearly households with less external contacts would be at the least risk from merging with others, and these policies should only be encouraged in regions where general social distancing has clearly reduced the prevalence of infection. Similar to our findings in earlier sections, our predictions are more optimistic when household and external contacts contribute less equally to transmission.

## Discussion

Here we show that the clinical and epidemiological features of COVID-19 interact to produce long expected delays between the implementation of strong social distancing measures and when their effects become apparent. Part of the delay is clinical. After infection, individuals generally pass through an asymptomatic incubation period before entering a phase of mild or moderate symptoms, and some fraction eventually require hospitalization. Documented deaths often occur after extended stays in critical care wards. The progression from initial infection to a reportable case (often at hospital admission) or death can be weeks, and is not interrupted by current interventions. In addition, social distancing measures reduce transmission outside the household, but in general they involve isolating individuals within their normal places of residence and thus do not prevent household transmission. They may in fact amplify it, by increasing the time household members spend together. If even a small fraction of households have been “seeded” with infection at the time an intervention is implemented, cases may continue to increase for multiple serial intervals. This residual transmission is exacerbated if weak inter-household connections remain, and especially if there are clusters of individuals less able to comply with social distancing measures, for example among communities with a high prevalence of “essential workers”.

Our results show that it is very difficult for interventions which only target transmission outside the house to effectively control the outbreak. Unless these interventions reduce the vast majority of contacts, ongoing transmission in households combined with occasional spillover to other households means that the epidemic may continue to increase long after social distancing begins and when it turns around, declines in cases can be extremely slow. We found that the relative contribution of household and external contacts to transmission was a critical determinant of the overall outcome of social distancing interventions, and the timescale over which effects could be observed. The number of contacts alone was not very informative for predicting intervention efficacy. It is not possible to predict the effect of an intervention that differentially affects household and external contacts by simply estimating the proportional reduction in the total R_0_. For example, even if the component of R_0_ from household transmission alone is greater than 1, infection cannot continue if external connections are substantially weakened. These findings highlight the need for more studies to determine the contribution of different types of contacts to transmission.

The role of household transmission in the spread of COVID-19 is variable across settings. Several studies with detailed contact tracing have attempted to estimate the household “secondary attack rate”, i.e. the probability of transmission per susceptible household member when there is a single infected individual in the house. In a large study in Shenzhen, China, Bi et al estimated this rate at 11% [34]. In Guangzhou, China the estimate was 20% [35], in Beijing 23% [36], in Zhuhai 32% [37], in Seoul, South Korea 16% [38] and in Taiwan, around 5% [39]. In a small German town with a large outbreak due to a superspreading event at a carnival, the household secondary attack rate was closer to 30% but decreased in larger households [40]. Liu et al considered a collection of known clusters involving close contacts in a single gathering (not just household, often group meals), and estimated a 35% secondary attack rate. Lewis et al find a rate of 28% in Wisconsin and Utah [41], while Grijalva et al found 53% in Wisconsin and Tennessee [42]. A recent review by Madewell et al [43] reports values between 4-44%. Curmei et al’s review [44] attempts to collect all these estimates and correct them upwards by accounting for false negative rates of diagnostic tests and for asymptomatic infections, resulting in estimates between ∼10-55%. Given that the average household size is relatively small in all these countries (∼3 or less), these numbers suggest that infection from outside the house must play a large role in order to explain the overall R_0_ values observed. By varying the relative weight of household vs external contacts, our study examined a range of household secondary attack rates from ∼10% to ∼65%. Many more studies have examined the role of household transmission in influenza spread, but the results are also equivocal: a review by Tsang et al found that household secondary attack rates varied from 1-40% across studies [45]. A massive cohort study from Japan recently shone some light on this complexity; finding that the risk of household influenza transmission was highly dependent on household structure and on the familial relationship between the primary and secondary case [46].

The networks we use to simulate infection were parameterized based on detailed surveys that used “contact diaries” to track the number of individuals someone interacted with on a randomly chosen day [20,21]. Contacts were generally defined as physical contact or face-to-face conversations, and were meant to capture interactions thought to be important for the spread of droplet-borne respiratory infections like influenza and coronaviruses. Like others, the data we use from these studies is the average number of daily contacts by age of each individual in the pair. However, these surveys also collected information on the duration and frequency of contacts, which could be used in the future to create dynamic networks with a more complex distribution of weights for each types of contact. One limitation of these sorts of surveys is that they are “ego-centric”, meaning that they only inform the distribution of the number of contacts but not the higher order network structure, which can be important for infection spread [26,47]. When we constructed our multi-layer network of external contacts, we used additional information from other studies to include clustering and modularity in our networks. Another limitation is that certain contacts that might be relevant to respiratory infections may be missed in surveys. For example, transmission via contaminated surfaces can occur between individuals who have never directly interacted, as can transmission in group settings where air is shared (e.g. in fitness classes [48] or at restaurants [49]).

There are multiple strategies to augment social distancing policies by reducing household spread, and these have been implemented to different degrees in different countries. We have not considered such combination policies in our analysis, but other models have explored them in detail. Household spread would be reduced by earlier diagnoses of cases (as soon as symptoms begin), proactive testing of exposed household members of cases, options for out-of-home care for individuals with mild symptoms, or better education and assistance with individuals caring for sick household members to avoid infection, for example via household use of face masks and disinfectants [36]. Population-level contact tracing initiatives would obviously also help [50,51]. Early and influential modeling studies that provided the impetus for widespread social distancing policies around the world assumed these policies would be accompanied by case-based interventions that would reduce household spread (e.g. [11,52]), but these measures have not been uniformly adopted, and are still completely absent in most of the United States.

Clearly a major determinant of the efficacy of social distancing policies for COVID-19 is the fractional reduction in contacts, but quantifying this value is difficult. A variety of data sources can provide some information. Surveys conducted in Wuhan and Shanghai, China comparing contacts before and after COVID-19 lockdowns found that the average number of daily contacts was reduced from ∼14 in Wuhan and ∼20 in Shanghai to ∼2, suggesting a more than ∼95% reduction in external contacts [53]. In the US, nationally-representative polls in late March/early April found that around three quarters of households were self-isolating [54], and estimated a mean reduction in contacts around 80% [55,56]. Since contact surveys are rare, measures of reductions in human mobility have been used as a proxy for contact rate reductions. Google [57] and Apple [58] provide reports on mobility changes based on user locations sourced from their smartphone mapping apps, as does Cuebeq [59]. Transit, a live-tracking and schedule-aggregating application for public transit, reports changes in service use [60], and SafeGraph publishes changes in foot traffic to different classes of locations [61].

Different measures of mobility often give very different estimates for the efficacy of social distancing interventions. For example, Klein et al found peak US national average reductions in both the radius of mobility and the number of events where device users came within near proximity of each other were about ∼50%, whereas communing volume was reduced by ∼75% [62,63]. For the same time period, Apple reported ∼50% reductions in direction requests, Transit reported ∼70% reduction in transit use, Google reported ∼40% reductions in visits to retail locations and ∼50% in visits to workplaces, and SafeGraph reported an ∼80% reduction in foot traffic at bars but only a 20% reduction to grocery stores. Together these results suggest that our simulations assuming ∼80% reduction in external contacts - which still often only results in mediocre outcomes - are likely overestimates, if anything, of reality. Wellenius et al attempted to infer the association between these mobility reductions and the particular social distancing policies that caused them. Using Google data they concluded that in the US, initial emergency declarations lead to ∼10% reductions, that each additional policy led to another ∼20% reduction, and that “shelter-in-place” orders resulted in additional ∼30% reductions [64]. By comparing mobility changes to estimates of R_0_ from case counts in countries around the world, Bergman et al estimated that each ∼10% reduction in mobility resulted in an ∼0.05 reduction in R_0_ [65]. Interestingly, they also found two other results in agreement with our findings here: there was a long delay between reductions in mobility and reductions in inferred R_0_ in many regions, and, the association between reductions in mobility and R_0_ was weaker in regions who implemented large scale contact tracing, which likely reduces household transmission.

Our results highlight the importance of residual contacts between households that remain despite social distancing measures. Many of these contacts are likely to be driven by individuals who must continue to work. Our own analysis of occupations held by residents of Philadelphia, USA, population ∼1.5 million, suggested that ∼30% of workers had jobs that fell into categories flagged as “essential”. A review by Lan et al of case reports within the first month of the outbreak in multiple countries found that about 15% of these cases were clearly work-related, and that even earlier in the outbreak, this was as high as 50% [66]. A report released by the UK Office of National Statistics found COVID-19 related deaths were much higher in certain occupational groups (e.g. relative risk of 4.5 for male security guards and 2 for female care workers), and the UK Biobank found an 8x higher rate of COVID-19 diagnoses in healthcare workers compared to the general population. Major clusters of infection have occurred in workplaces as varied as call centers [38] and meatpacking plants [67]. The results of our modeling show how risks to essential workers spill over to others, increasing the individual infection risk for workers’ household members and increasing the persistence time of epidemics in the community at large. Given the limited apparent ability to reduce workplace contacts and transmission, reducing household transmission or other external contacts may be even more important. Another strategy we have not considered is selective restructuring of contact networks to increase clustering and decrease mean path length, so that transmission risk is minimized without further reducing contacts [68].

Separate assumptions of our modeling approach could lead our predictions to be slightly pessimistic. We assume a baseline value of R_0_ ∼ 3, whereas some other studies have used values between 1.9 - 2.7 [4–6,51,69]. There are several reasons why we believe those estimates are likely a little too low. Firstly, they tended to assume very short serial intervals and infectious periods, whereas other studies have estimated longer serial intervals [34,70–72], particularly in the absence of quick isolation of mild cases, which is more likely to reflect what is going on in most of the world outside of east Asia. Secondly, those estimates often fit to cases counts that were doubling every 5-6 days, whereas in many settings doubling times were closer to 3 days early in the outbreak [73–76]. Finally, nearly all previous estimates of R_0_ fit a randomly-mixing population (with or without age structure), whereas in our highly structured network population, higher R_0_ values are needed to achieve the same doubling time. R_0_ values as high as 3-6 have been estimated using rigorous model-fitting methods [9,52,72]. With lower R_0_ values, any estimates of the % reduction in external contacts needed to achieve a certain rate of reduction of cases and deaths would be reduced. However, our main qualitative results about delays to epidemic peak and the complex role of household transmission hold.

Our results are not sensitive to our assumptions about the fraction of cases that progress to more serious clinical stages nor to the case fatality risk. However, our estimates for the timing of peak values do depend on the distribution of delays we assume, for example between symptom onset and hospitalization, or between ICU admission and death. There is variability in the estimates of these values across studies (see Supplementary Methods), and these values likely differ by country, depending on the standard of care and the underlying health of the population. While we have considered wide intervals for the interpatient variation in these durations, we have not propagated uncertainty in the distribution of these values. We hope that by providing our code, researchers who are interested in specific contexts where these values may differ significantly can explore those scenarios. There are other factors which influence the delay between implementation of social distancing measures and peak cases and deaths that we have not included in our model. One factor is reporting delays, which may be especially long for deaths in certain regions. Another factor is that there could be a delay between implementation of distancing measures and adoption by a majority of the population.

By including more details of transmission network structure, we are able to examine effects that would not be apparent in well-mixed epidemic models. However, our population structure is still simplistic in many senses. For example, we do not explicitly model the dynamics of certain institutions that have been particularly hard-hit by COVID-19, such as retirement homes and long-term care facilities [77], prisons [78,79], and homeless shelters [80]. Understanding the unique contact networks, transmission risks, host susceptibility, and mortality risks in these populations is an important area for future research. We also do not consider the potential for hospital-acquired transmission and the role of healthcare workers. Doctors, nurses, and other health professionals are reported to make up 5-10% of cases in some regions, and while increased testing is likely one factor driving these rates, it is clear that there are also unique risks to this profession.

Strong social distancing measures tend to be economically costly and in most regions of the world these measures were relaxed to some extent after a few months. In this study we examined a particular partial relaxation strategy in which households form “bubbles” with other households. We predicted that widespread adoption of these bubbles should only occur in the context of decreasing incidence and compensatory reductions in external contacts, in order to maintain epidemic control. Here we imagine that household bubbles are formed voluntarily for social reasons, but households may also be forced to “double-up” when one household experiences eviction from their current residence. The economic recession in the US has led to massive increases in households at risk of eviction, and separate work using a similar model found that evictions could result in substantial increases in cases across cities if the current eviction bans expire [81]. Other modeling studies have explored the impact of generalized relaxation of social distancing on second-wave scenarios [82–85]. Although not the primary focus of this work, when we simulate generalized relaxation we find that cases always begin to increase almost immediately. This is in contrast to the months-long delays between relaxation of interventions and resurgences of cases observed in many parts of Europe and North America in summer through fall 2020. In reality other factors not included in our model are likely to play a role in observed delays post-relaxation, such as a delayed behavioral response to relaxation policies, shifting age distributions of cases, repeated stochastic re-introductions and extinctions, and seasonality. We did observe variable delays until deaths and hospitalizations began to increase again in our simulations, which was explained by the clinical progression times and the degree of relaxation (Figure S7).

Many studies are now attempting to estimate the degree to which different social distancing measures (e.g. school closures, stay-at-home policies) reduce the reproductive ratio or the exponential growth rate of cases. Our results point out a few challenges to these efforts. The long delays we describe in this paper mean that methods that fit simple growth functions to data and look for changes in their values may have trouble identifying effects. If there are a series of interventions that tend to be implemented in similar orders or at similar intervals across settings, and the goal is to estimate the effect of each (e.g. [11,86]), then the delays we describe here could lead to falsely attributing the effect of one intervention to another that occurs later (e.g. see [8] and comments in response). Some of these problems can be avoided by explicit use of mathematical models that take into account the prolonged clinical progression of COVID-19 (e.g. [12,87]), which is the first order cause of these delays. However, our results show that transmission network structure also plays an important role. Importantly, the amount by which overall transmission is reduced by social distancing measures and the delay until effects are seen depends on the relative role of household vs external transmission, which is unknown and may be different by setting.

## Data Availability

We provide all of our code online. The differential equation version of our model is available as an online simulation tool and our stochastic network implementation code is open source and hosted on Github.

https://alhill.shinyapps.io/COVID19seir/

https://github.com/alsnhll/COVID19NetworkSimulations

## Acknowledgments

This work was supported by grants from the US National Institutes of Health DP5OD019851 (ALH), R01AI146129 (MZL), and R01AI101229 (MZL). We thank Julianna Shinnick for help in data collection and attendees at the HIV Dynamics & Evolution Conference for helpful feedback.

## Supplementary Methods

### Model of COVID-19 spread and clinical progression

We use a compartmental epidemiological model, based on the classic SEIR model, to describe the spread and clinical progression of COVID-19 (**Figure 2**). The basic model structure is inspired by many studies on the natural clinical progression of COVID-19 infection. Infected individuals do not immediately develop severe symptoms, but instead pass through milder phases of infection first. For a nice summary, see Wu and McGoogan [88]. Susceptible (S) individuals who become infected start out in an exposed class E, where they are asymptomatic and do not transmit infection. Progression from the exposed stage to the infected stage I, where the individual is symptomatic and infectious, occurs after an average duration of *T*_*E*_days. The clinical descriptions of the different stages of infection are given below. Infected individuals begin with *mild* infection (I_1_) that lasts on average *T*_*1*_ days, from which a fraction (*f*_1_)progress to *severe* infection (I_2_) as opposed to recovering. Severe infection progresses to a *critical* stage (I_3_) in a portion (e*f*_*2*_) of individuals and resolves in the rest, after an average duration *T*_*2*_. A fraction (*f*_*3*_) of individuals with critical infection die, while the remainder recover. The average duration of critical infection is *T*_*3*_ days. Recovered individuals are tracked by class R and are assumed to be protected from re-infection for the duration of the epidemic. Individuals may transmit the infection at any stage, though with different rates. The transmission rate in stage *i*, β_i_.

The clinical definitions of the infection stages are as follows

- Mild infection - These individuals have symptoms like fever and cough and may have mild pneumonia. Hospitalization is not required (though in many countries such individuals are also hospitalized)
- Severe infection - These individuals have more severe pneumonia that leads to dyspnea, respiratory frequency <30/min, blood oxygen saturation <93%, partial pressure of arterial oxygen to fraction of inspired oxygen ratio <300, and/or lung infiltrates >50% within 24 to 48 hours. Hospitalization and supplemental oxygen are generally required.
- Critical infection - These individuals experience respiratory failure, septic shock, and/or multiple organ dysfunction or failure. Treatment in an ICU, often with mechanical ventilation, is required.

A summary of the variable definitions:

- S: Susceptible individuals
- E: Exposed individuals - infected but not yet infectious or symptomatic
- I_i_: Infected individuals in severity class *i*. Severity increases with *i* and we assume individuals must pass through all previous classes
  - I_1_: Mild infection
  - I_2_: Severe infection
  - I_3_: Critical infection
- R: Individuals who have recovered from disease and are now immune
- D: Dead individuals
- N=S+E+I_1_+I_2_+I_3_+R+D Total population size (constant)

To describe the average time course in a large, well-mixed population, the model can be represented by the following set of differential equations:

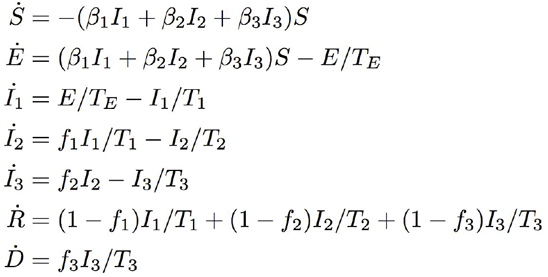

The basic reproductive ratio R_0_ of this model is:

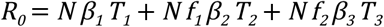

Our model makes several assumptions with regards to the clinical and epidemiological dynamics of COVID-19. We do not explicitly track asymptomatic infections, which have been estimated to be around 20% of infections [34] (but estimates range from 1% to 50%, reviewed in [89]). However, in our model asymptomatic infections can be considered to be part of the “mild infections” compartment. There is significant evidence that asymptomatic individuals are infectious, and with this assumption their infectiousness would be equal to those with mild infection. For simplicity we assume that critical infection always occurs after passing through a stage of severe infection, while in reality there appears to be some individuals who progress directly from mild to critical infection. Similarly, we assume that only individuals who have progressed to critical infection die, though there is now some evidence that death can occur unexpectedly in individuals who are not already hospitalized. We do not think that any of these assumptions affect the main conclusions of our paper, which does not focus on hospital resource use or morbidity/mortality estimates. In general our model allows for individuals at all stages of infection to transmit to others. However, for this paper we have assumed that only individuals with mild infection can transmit. We think it’s likely that an individual is most infectious during this stage, when they would still be in the community and feeling well enough to interact with others. We thus ignore transmission from hospitalized patients to their healthcare providers (*β*_2_=*β*_3_=0). We make the standard SEIR model assumption that the time until an infected individual experiences symptoms is the same as the time until they become infectious, whereas it has now been shown for COVID-19 that there is significant transmission risk starting ∼1 day before symptom onset. While this disease feature is important for studies that examine symptom based case detection/isolation, for simplicity we have ignored this feature in our model, which doesn’t focus on these issues. The serial interval produced by our model parameters agrees with observed values, and so we believe our timescales for infections spread are realistic. We do not explicitly consider age-dependent rates of infection or progression to more serious stages.

The differential equation version of our model is available as an online simulation tool in which users can explore the effects of these assumptions of model structure as well as parameter values on model outputs: https://alhill.shinyapps.io/COVID19seir/

### Stochastic network implementation

To avoid the assumptions inherent in formulating an epidemiological model using standard differential equations (e.g. assuming a randomly-mixing population, deterministic dynamics, and exponentially-distributed durations of each infection stage), we implement the model as a stochastic process simulated on a transmission network.

We believe that this implementation is important for our research questions, for a few main reasons. Firstly, respiratory infections are spread by close physical contact that occurs in highly structured contact networks, which impact the growth rate of outbreaks, the relationship between an individual’s inherent infectiousness and the basic reproductive ratio R_0_, the variability in secondary infections between individuals, and the effects of control measures on spread. Secondly, typical differential equation models implicitly assume that the durations of infection stages are extremely long-tailed, which can lead to unrealistic conclusions about the timescale of the epidemic response to control measures as well as to the relationship between the early growth rate, serial interval or generation time, and R_0_. Finally, we believe it is important to model the uncertainty in epidemic trajectories that arises from the inherent stochasticity of transmission.

In our stochastic formulation, we assumed that the duration of each stage of infection was gamma-distributed, with both the mean and variance taken from the literature. Individuals were connected with a fixed, weighted contact network that determined the potential paths of transmission. The network is represented as a sparse matrix to save memory (a list of the index of each node in an edge, along with the edge weight). The model was implemented with a discrete time stochastic process that tracked the state of each individual and the time since they first entered that state. The model was implemented in Python using JAX, a framework for generating high-performance code optimized to run on GPUs. Our code was entirely run in the cloud on Google Colab. The code is available at : https://github.com/alsnhll/COVID19NetworkSimulations

### Model parameters

We estimated the distribution of the duration of each stage of infection from the literature. For the incubation period, we used an estimate of 5 ± 4 days from [34,90], which is consistent with most other estimates. The duration of mild infection, which we assume is roughly equal to the infectious period, can be estimated from a few different sources: a) the duration of mild symptoms, b) the time from symptom onset to hospitalization (e.g. time to progress to severe stage), or c) the duration of viral shedding via sputum or throat swabs, d) inferred from both the incubation period and the “serial interval” between symptom onset in an index case and a secondary case they infect. Considering (d), we took an estimate of the serial interval from Bi et al [34], who used a large cohort of transmission pairs and reported values separately for situations where the index case was not rapidly isolated or hospitalized during mild infection. Similar values were found in [69–72]. Some other studies have estimated smaller values (e.g. [51]), likely due to rapid isolation of symptomatic cases in Asia [70]. This value implies an infectious period of 6 ± 2 days. This is relatively consistent with estimates of the time from symptom onset to hospitalization [52,91–93], and is roughly equivalent to the time at which average viral loads drop 4-5 orders of magnitude from peak values [94].

The duration of severe infection in our model is equivalent to the time from hospital admission to recovery for individuals who did not progress to the critical stage, or the time from hospital admission to ICU admission (since critical cases require ICU-level care). Since it is hard to find direct estimates of this duration, we instead used estimates of the total time from symptom onset to ICU-admission (e.g. combined length of mild + severe infection) [92,93], and subtracted the inferred duration of mild infection. This led to an estimate of 6 ± 4.5 days. We estimated the duration of critical infection (the length of ICU stay) directly from cohort studies to be 8 +/-6 days [92,95]. With these estimates, we verified that the total time from symptom onset to death/recovery agreed with data (∼20 ± 10 days) [69,92,96]. Note that we assumed that the duration of each stage of infection was independent of the previous stage and was also independent of the eventual outcome.

In each stage of infection, an individual can either progress to the next stage or recover, and the probability of each was also estimated from the literature [8,88,97]. These large clinical cohort studies estimated that ∼ 20% of infections required hospitalization (i.e. progress to severe stage), ∼5% require ICU care (i.e. progress to critical stage), and an overall case fatality risk of 2%. This leads to probabilities of progression of 0.2 from I_1_ → I_2_, 0.25 from I_2_→ I_3_, and 0.4 from I_3_ → D. However, there is significant uncertainty in these values. More recent estimates suggest the rates of progression to more serious stages of infection might be lower after correcting for asymptomatic/undiagnosed cases in China [96], but in contrast, recent studies from the US suggest higher rates of ICU admission and death among hospitalized patients compared to in Asia [98,99]. Since the results of our paper focus on the timing of the peaks of different infection stages but not on the prevalence at these peaks, they are not sensitive to these assumptions.

We chose a value of the transmission rate β such that the early epidemic doubling time was ∼ 4 days (growth rate r = 0.17/day) and the basic reproductive rate R_0_ = 3. While some studies have estimated R_0_ values as low as 2, these have generally used estimates based only on the serial interval which are mathematically problematic, used short estimates of the serial interval that are influenced by rapid case isolation in Asia as opposed to longer values found in other studies that correct for this, and fit to longer doubling times that what was observed early in the outbreak in the US and Europe. For a simple unweighted network, we estimate β using the formula R_0_ = *βT*_*1*_ (*n* - 1) where, *n* is the mean degree of the network and *T*_*1*_ is the average duration of mild infections. This relationship is modified for the weighted two-layer network,

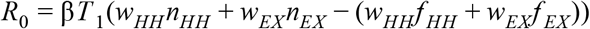

where, 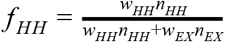 and *f* _*EX*_ = 1 − *f* _*HH*_ with the weight and mean degree of the household (external) layer given by *w*_*HH*_ (*w*_*EX*_) and *n*_*HH*_ (*n*_*EX*_). As this equation suggests, the rate of infection is constant per time per contact for a given contact type, and hence would be described as “density-dependent” transmission.

The time at which we implemented interventions in our simulations was chosen to occur at a realistic time in the epidemic relative to real responses to COVID-19. To estimate this time for a typical US metro area, we downloaded case and death counts for each US county and aggregated them into the top 50 largest metro areas. Then we collected the time that stay-at-home orders were implemented in each of these regions, and looked at the median cumulative case and death counts by that day, and chose our intervention time to match those values. We found that there were on average around 200 cases per million inhabitants at time of stay at home intervention (range [30,1000]), and ∼5 [0-20] deaths per million. We assumed that reported “cases” are hospitalized individuals only (cumulative I_2_). These values were recreated in our simulation in well-mixed populations on Day 40, which we used as the intervention time. At this time, ∼0.5% of the population had ever been infected (including E, I_1_, I_2_, I_3_, R, and D). Since the median day shelter-in-place orders were implemented across these metros was April 1, Day 0 in our simulation corresponds to ∼ Feb 20. For simulations in other networks, we kept R_0_ constant but due to the population structure, the exponential growth rate of the epidemic varied between different values of the household and external contact structure. Thus for each structure we chose slightly different intervention start times to keep the infection prevalence consistent across comparisons. The start times varied from Day 43 (when *w*_*EX*_ /*w*_*HH*_ = 3, 1), Day 45 (when *w*_*EX*_ /*w*_*HH*_ = 1/3) to Day 55 (when *w*_*EX*_ /*w*_*HH*_ = 1/9).

**Table 1:**
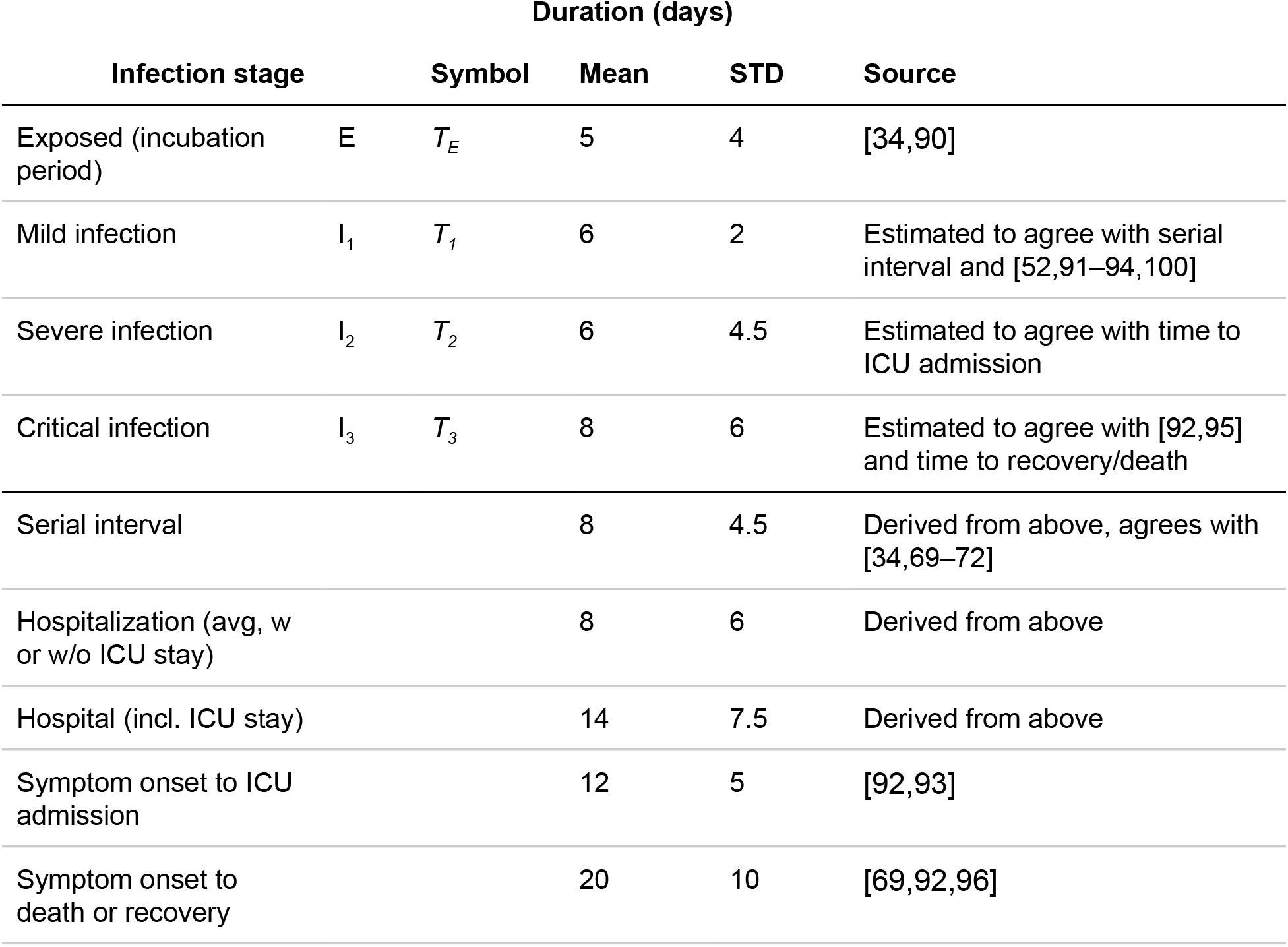
Model parameters for duration of each stage of infection.

| Duration (days) |  |  |  |  |  |
| --- | --- | --- | --- | --- | --- |
| Infection stage |  | Symbol | Mean | STD | Source |
| Exposed (incubation period) | E | $T_E$ | 5 | 4 | [34,90] |
| Mild infection | $I_1$ | $T_1$ | 6 | 2 | Estimated to agree with serial interval and [52,91–94,100] |
| Severe infection | $I_2$ | $T_2$ | 6 | 4.5 | Estimated to agree with time to ICU admission |
| Critical infection | $I_3$ | $T_3$ | 8 | 6 | Estimated to agree with [92,95] and time to recovery/death |
| Serial interval |  |  | 8 | 4.5 | Derived from above, agrees with [34,69–72] |
| Hospitalization (avg, w or w/o ICU stay) |  |  | 8 | 6 | Derived from above |
| Hospital (incl. ICU stay) |  |  | 14 | 7.5 | Derived from above |
| Symptom onset to ICU admission |  |  | 12 | 5 | [92,93] |
| Symptom onset to death or recovery |  |  | 20 | 10 | [69,92,96] |

**Table 2:** Model parameters for probability of progressing from each stage of infection.

|  |  | Probability of progression |  |  |
| --- | --- | --- | --- | --- |
| Infection stage |  | Symbol | Value | Source |
| Exposed (incubation period) | E |  | 1 |  |
| Mild infection | $I_1$ | $f_1$ | 0.2 | [8,88,97] |
| Severe infection | $I_2$ | $f_2$ | 0.25 | [8,88,97] |
| Critical infection | $I_3$ | $f_3$ | 0.4 | [8,88,97] |

### Well-mixed network structure

We approximated a well-mixed population by randomly connecting each individual to 100 other individuals in the population. While a truly “well-mixed” network would connect every one to everyone else, with the large population sizes we use (10^6^), this would require huge amounts of memory and negate the computational efficiency of using sparse matrices to represent the networks. Since in reality each individual will only transmit to a few others before recovering, any uniform random network with a large degree is a good approximation to a fully-connected network.

### Two-layer household network structure

#### Construction

We constructed a weighted two-layer network, consisting of a layer for within-household connections and another for external connections. Individuals were first assigned households using the distribution of household sizes in the United States (data obtained from the 2010 census, mean household size *n*_*HH*_ ∼ 2.5). All individuals in a household were connected to each other. Each individual was then assigned random external contacts. The degree distribution for external connections was obtained from contact survey data that recorded daily interactions of individuals [20,21]. This data was originally collected for a subset of countries and then projected to other countries using a wide range of demographic data. The contacts are age structured and include the type, duration, location and frequency of the contact. These surveys were designed to capture interactions relevant to the spread of respiratory diseases, and include interactions consisting of either physical contact or close face-to-face conversations. From this data, we obtained daily non-household contacts by summing ‘work’, ‘school’ and ‘other’ contacts. For the United States there were on average *n*_*EX*_ ∼ 7.5 ± 2 (mean ± standard deviation) daily non-household contacts, equivalent to 3x household contacts. The variance was obtained by the variance in the mean across all age groups. We assigned external degrees to individuals following a binomial distribution with this mean and standard deviation. This assignment was done randomly and did not depend upon the size of the person’s household. The network layer was created by giving ‘stubs’ to individuals equal to their external degrees. These stubs were then randomly paired between individuals in the population to create connections. In addition to the number of contacts, the probability of infection also depends upon the duration and intensity of the contact. We assigned weights (*w*_*HH*_, *w*_*EX*_) to the two layers of the network to account for this. We considered different scenarios (keeping R_0_ fixed) where, *w*_*HH*_ = and varied *w*_*EX*_ = 3, 1, 1/3, 1/9 to reflect the relative importance between household and external contacts.

In order to investigate the robustness of our results with respect to details in the large-scale clustering of the network, we also constructed a hierarchically structured external layer using ideas from nested metapopulation models [101,102]. Individuals were assigned to ‘neighborhoods’ of size 10,000 (there are 100 such neighborhoods for the population size of 1 million) and as an extreme case 90% of their external connections were forced to be within the same neighborhood with *n*_*EX*_ ∼ 7.5 ± as before.

#### Intervention

Intervention, corresponding to social distancing measures, was modeled by reducing the weight of the external layer, *w’*_*EX*_ = (1 − ε) *w*_*EX*_ where, ε (0 ≤ ε ≤ 1) is the ‘intervention efficacy’. When we are in the parameter regime where the probability an individual infects any given external contact over the duration of their infectious period is significantly less than one (i.e. R_0_ ^EX^ < *n*_*EX*_), reducing the weight of external contacts is equivalent to randomly removing external contacts (i.e. *n’*_*EX*_ = (1 − ε) *n*_*EX*_). Only in the regime where the transmission rate per contact is so high that R_0_ ^EX^ is limited only by the number of contacts would these two alternative methods of implementing social distancing lead to different results. Household contacts were either unaffected (*w’*_*HH*_ = *w*_*HH*_) or doubled (*w’*_*HH*_ = 2 *w*_*HH*_) during intervention, to represent the increased time spent with household members.

#### Partial relaxation of intervention

We modeled a scenario of partial relaxation of social distancing measures in which every household could “merge” with another household. This relaxation altered the household layer so that all households in the network were paired with another random household and the two were merged to create a fully-connected joint household (*n’*_*HH*_ ∼ 5). We considered two scenarios for how the external layer changed during partial relaxation of intervention. In the baseline scenario, the external layer was the same as during intervention. In a second scenario, the number of external contacts per individual was reduced so that the average number of total contacts was the same before and after merging (*n’*_*EX*_ = *n*_*EX*_ *- n*_*HH*_). This partial relaxation was only conducted for the scenario where household and external connections had the same weight (*w*_*HH*_ = 1 and *w*_*EX*_ = 1).

#### Individual probability of infection as a function of household size

We computed the individual probability of infection retrospectively, after the epidemic had died out, for the case of a 100% effective intervention. We counted the number of individuals who were ever infected by the end of the epidemic and calculated the corresponding distribution of their household sizes. This is a conditional probability that gives the probability of household size, given that an individual was infected P(HH size|inf). The probability of infection given household size was then calculated using Bayes’ rule, P(inf|HH size) = P(HH size|inf) * P(inf)/P(HH size) where, P(inf) is the fraction of the total population that was infected during the epidemic and P(HH size) is the distribution of the household sizes in the population. The probability was averaged over 10 iterations of the simulation.

### Five-layer network structure

We constructed a non-weighted 5-layer (1 household and 4 external) network for an age-structured population with realistically structured external layers. The population was divided into four age groups: preschool aged (ages 0-5), school-aged (ages 5-19), working-aged (ages 19-64) and elderly (ages 65+). The distribution of ages in the United States was obtained from the 2018 World Bank Age Structure data which consists of the population divided into 5-year age groups. In addition to the broad age groups, we also kept track of the 5-year age group that an individual belonged to. The contact survey data [21] used to construct the 2-layer network was also predominantly used here to determine the statistical properties of the layers.

#### Household layer

The household layer was constructed in the same manner as in the 2-layer network. Individuals were assigned to fully-connected households using the distribution of household sizes in the United States from the 2010 census data (mean household size, *n*_*HH*_ ∼ 2.5).

#### School layer

The school layer contained connections between the school-aged age group in the population. This external layer was constructed using the technique described by Ball et al [103], where a network layer with a given degree distribution and correlations among connections (i.e. “clustering” or “transitivity”) can be constructed. The degree distribution in this layer was obtained from the school contact survey data [21], which estimated that the school-going population in the United States, on average, had *n*_*SCHOOL*_ ∼ 7.3 ± 1.8 school-related contacts daily. The variance was obtained by the variance in the mean across the school-aged (5-19) age groups. Individuals were assigned degrees randomly from a binomial distribution with this mean and standard deviation. The age-structure of contacts at school suggests a significant level of clustering by age group. To create correlated connections, the method of Ball et al requires two additional parameters: The number of groups *n* to divide the population into, and *r*, which is roughly the extent to which individuals within the same group are connected. *r* = 1 corresponds to individuals being connected only within their group whereas *r* = 0 implies completely random connections without any correlation with members of their own group. Averaged over the school age groups (5-19), we found that ∼ 57% of school contacts for an individual belonged to their own age group. Since there are three 5-year age groups in our school-aged population, we divided our population into those three groups (*n* = 3) and chose *r* = 0.57.

#### Work layer

The work layer consisted of connections between the working-aged group in the population. This layer was constructed similarly to the school layer. We obtained the mean and standard deviation in number of daily work contacts from the contact survey data (5.0 ± 2.3).

The variance was obtained by the variance in the mean across working-aged (20-64) age groups. Individuals were randomly assigned a work-place degree from a binomial distribution with the mean and standard deviation. Then we used a separate study by Potter et al that mapped real work-place networks to estimate the level of clustering (transitivity), at ∼0.1 [15]. Note that Potter et estimated a higher degree, since their network contained *total* contacts in a work network and not just the daily contacts as given by the contact survey data, but the coefficient of variation is similar. We again used the Ball et al technique [103] to create clustering in the network. High transitivity can be achieved by choosing appropriate correlation parameters *n* and *r*. We assumed this clustering arose from the fact that people belonging to the same work-place have a higher chance of being connected to each other. According to NAICS (North American Industry Classification System), ∼ 80% of businesses in the United States have ≤ 10 employees and so for the sake of simplicity, we assigned each individual to a ‘work-place’ of size ten 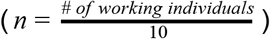. We picked *r* = 0.8 in order to have a high level of transitivity (∼ 0.12) while also ensuring inter work-place connections.

#### Friend (social contacts) layer

The friend layer represents social contacts of people, their ‘friend circles’. Individuals were assigned to fully-connected friend groups, where friend groups consisted of people belonging to the same age group (preschool, school, working, elderly). The distribution of sizes of these friend groups were obtained from Wrzus et al [27], which is a meta-analysis of the effects of age on social networks. From their data, we estimated that the mean sizes of friend groups for individuals less than 20 years of age is ∼ 10, that for the working age population ∼ 7 and ∼ 5 for the elderly. These constitute *total* social contacts for the individuals and not just their daily interactions as given by the contact survey data. The ‘other’ category in the contact survey data [21] gives the number of daily interactions that an individual has that are not within the household, at school or at the work-place. In our network, the friend layer and the community layer together constitute this ‘other’ category. According to the data, averaged over the age groups, there are ∼ 4.3 ± 1.9 such interactions for an individual in the United States. The variance was obtained by the variance in the mean across all age groups. Since the total social contacts are much higher than this daily value, we scaled the sizes of the friend groups (*n*_*FRIEND*_ → *n*_*FRIEND*_ /4) to reflect daily social contacts. The scaling factor was chosen to ensure that the community layer didn’t end up being too sparse. We chose to do this instead of creating a weighted network for the sake of simplicity. The friend group sizes were drawn using the negative binomial distribution to account for the large variance (estimated variance post scaling: 6.25, 2.5, 1.75 for the three age-groups) seen in the data.

#### Community layer

This layer constitutes the additional random contacts an individual has during the course of their day. As mentioned previously, together with the social layer this constitutes the ‘other’ category in the contact survey data. We chose the average degree for this layer, *n*_*COMM*_ ∼ 1.72 ± 0.76 such that the combined degree for the two layers matches what is seen in the data. We used a binomial distribution and assigned degrees randomly to the individuals. This layer was constructed in the same manner as the external layer in the 2-layer network. It amounts to using the Ball et al [103] technique with *n* = 1 and *r* = 0.

#### Sensitivity analysis

We introduced the idea of “neighborhood” clustering to check the robustness of the results to clustering of contacts at an intermediate-scale between households and the city as a whole. As in the 2 layer network, individuals were assigned to neighborhoods of size 10,000. School and community layer contacts for individuals were predominantly within their neighborhood. The community layer was created by enforcing 90% (*r* = 0.9) of the connections to be within the same neighborhood (*n* = 100) with *n*_*COMM*_ ∼ 1.72 ± 0.76. School-aged individuals from each neighborhood, belonging to the same 5 year age group, were assigned to schools of size 500 (∼ average size of public schools in the US according to the 2017 data from the National Center for Education Statistics). The degree distribution of the school contacts was the same as before, *n*_*SCHOOL*_ ∼ 7.3 ± 1.8 but all school contacts for an individual were restricted to their own school 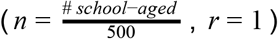.

#### Intervention

The social distancing intervention was modeled by deleting a certain percentage of external connections. School connections were always completely deleted during intervention, whereas social and community connections were deleted depending upon what we termed as the ‘efficacy’ of intervention. For example, 85% efficacy corresponds to deleting 85% of contacts in these layers. We considered two scenarios for the effect of intervention on the work-layer. In one case, similar to the social and community layer, a percentage of work contacts was deleted depending upon the intervention efficacy. Random deletion of connections leads to a large reduction in the transitivity of the work layer, from ∼ 0.12 to ∼ 0.02. In the second case, the effect of intervention was modeled as a clustered deletion of workplaces instead. For example, 85% intervention efficacy corresponds to 85% of workplaces being dissolved. Clustered deletion still maintains a high level of transitivity ∼ 0.10 in the work layer. Note as all school connections were completely deleted during intervention, the total effective efficacy of intervention was a bit higher than that for the social, community and work layers. For example, 85% efficacy in these external layers corresponds to a total efficacy of 88%.

#### Probability of infection for an individual working during intervention

We calculated the probability of infection for two intervention efficacies (85%, 70%) at day 300 after the start of the outbreak. We first obtained the fraction of the infected people (at day 300) who were still working during the intervention. This gives the conditional probability that an individual was working during intervention given that they were infected, P(working|inf). We calculated the probability of infection given that an individual was working during intervention using Bayes’ rule, P(inf|working) = P(working|inf) * P(inf)/P(working) where, P(inf) is the fraction of the total population that was infected by day 300 of the epidemic and P(working) is the fraction of the total population that was working during intervention. The probability was averaged over 10 iterations of the simulation.

#### Probability of infection for an individual living with working household members during intervention

This was also calculated at day 300 of the epidemic for the two intervention efficacies and, averaged over 10 iterations of the simulation. We obtained the fraction of the infected people (at day 300) who had at least 1 household member who was working during intervention. This corresponds to the probability that an infected individual had at least one working household member during intervention, P(HH working|inf). As before, the probability of infection given that an individual was living with working household members during intervention was then calculated using Bayes’ rule, P(inf| HH working) = P(HH working|inf) * P(inf)/P(HH working) where, P(inf) is the fraction of the total population that was infected by day 300 of the epidemic and P(HH working) is the fraction of the total population that had at least one working household member during the intervention.

#### Probability of infection for a non-working individual with no working household members during intervention

This was calculated in the same way as the other probabilities. We obtained the fraction of the infected people (at day 300) who were not working and had no working household member, P(HH not working|inf). As before, the required probability was then calculated using Bayes’ rule, P(inf| HH not working) = P(HH not working|inf) * P(inf)/P(HH not working) where, P(inf) is the fraction of the total population that was infected by day 300 of the epidemic and P(HH not working) is the fraction of the total population that was not working and had no working household members during the epidemic.

### COVID-19 Data

#### Wuhan, China

Data from Wuhan, China was obtained from the two daily situation reports published each day: http://wjw.wuhan.gov.cn/ztzl_28/fk/yqtb/index_11.shtml. We extracted new cases, new deaths, the number of individuals currently hospitalized with severe infection (which we took as a proxy for “hospitalized”) and the number of individuals currently hospitalized with “critical” infection (which we took as a proxy for individuals in the ICU). Data was already digitized by Li et al [104] for January and February (https://github.com/c2-d2/COVID-19-wuhan-guangzhou-data); we added data into March. For some dates the numbers of severe and critical cases were only reported for the entire province of Hubei, and in those instances, following Li et al, we assumed the proportion of those attributable to the city of Wuhan was equal to the proportion of currently active cases occurring in Wuhan. The vast majority of the Hubei outbreak took place in Wuhan.

#### Lombardy, Italy

Data on daily new cases and deaths as well as current patients hospitalized or in ICU for the Lombardy region of Italy was downloaded from Github repository maintained by the Department of Civil Protection in Italy : https://github.com/pcm-dpc/COVID-19`

#### Madrid, Spain

Data for daily new cases, daily deaths, daily new hospital admissions, and daily new ICU admissions were obtained from an online application maintained by the National Center for Epidemiology, using data from the Ministry of Health: https://cnecovid.isciii.es/covid19/. In the “documentation and data” section of the application there is a link to a .csv file with all the data used in the web app.

#### New York City, New York, USA

Daily new cases and deaths were downloaded from the Github repository of the New York City Department of Health and Mental Hygiene: https://github.com/nychealth/coronavirus-data. Number of currently individuals hospitalized or in ICU by date was obtained from the Github repository for digital news platform The City, who obtains this data directly from the New York state governor’s office: https://github.com/thecityny/covid-19-nyc-data

#### Los Angeles County, California, USA

Daily new cases and deaths were obtained from the Github repository maintained by the Los Angeles Times newspaper: https://github.com/datadesk/california-coronavirus-data. The number of individuals currently in the hospital or ICU was obtained from the California Health & Human Services Open Data Portal: https://data.chhs.ca.gov/dataset/california-covid-19-hospital-data-and-case-statistics/resource/6cd8d424-dfaa-4bdd-9410-a3d656e1176e?view_id=b23b0158-a85d-4bf2-95b1-96f7556f7342

## Supplementary Figures

**Figure S1:**
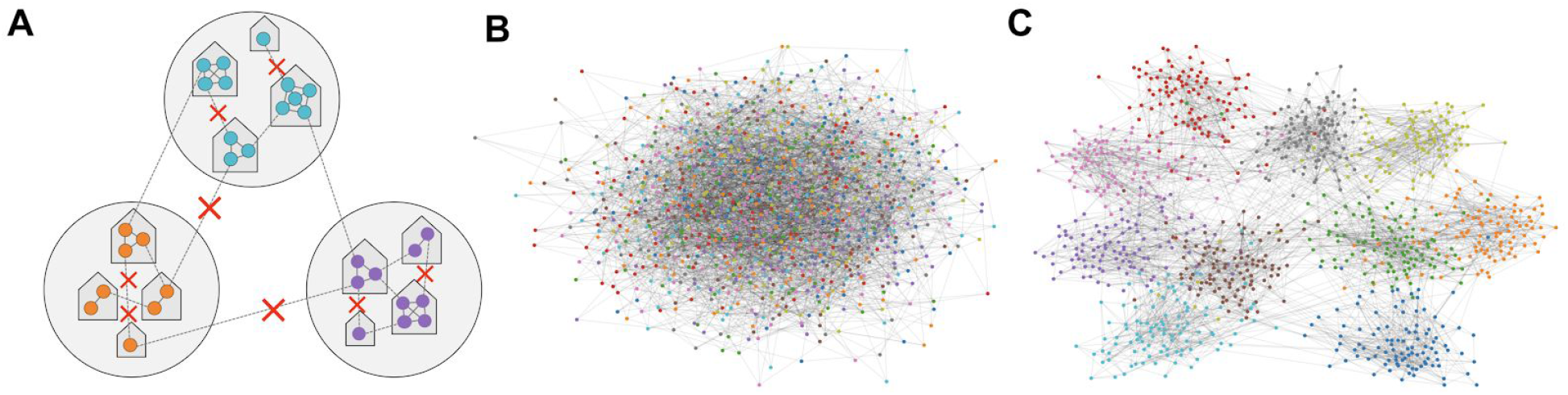
Creating hierarchical structuring in transmission networks via “neighborhoods”. A) Schematic of the construction of the networks neighborhood clustering. Each household belongs to a mutually-exclusive “neighborhood”, and external connections are preferentially created within the same neighborhood. B-C) Visualizations of connections in the external layer of the two-layer transmission network with and without neighborhood clustering. For ease of visualization, this is plotted for a population size of 1000 with 10 “neighborhoods” (node color) of size 100. In the real simulations, the population size is 10^6^ with 100 neighborhoods of size 10,000. A) The external layer consists of random connections between individuals, irrespective of their neighborhoods. B) 90% of external connections are preferentially attached to individuals within their own neighborhood, creating more intermediate-scale clustering in the network.

**Figure S2.**
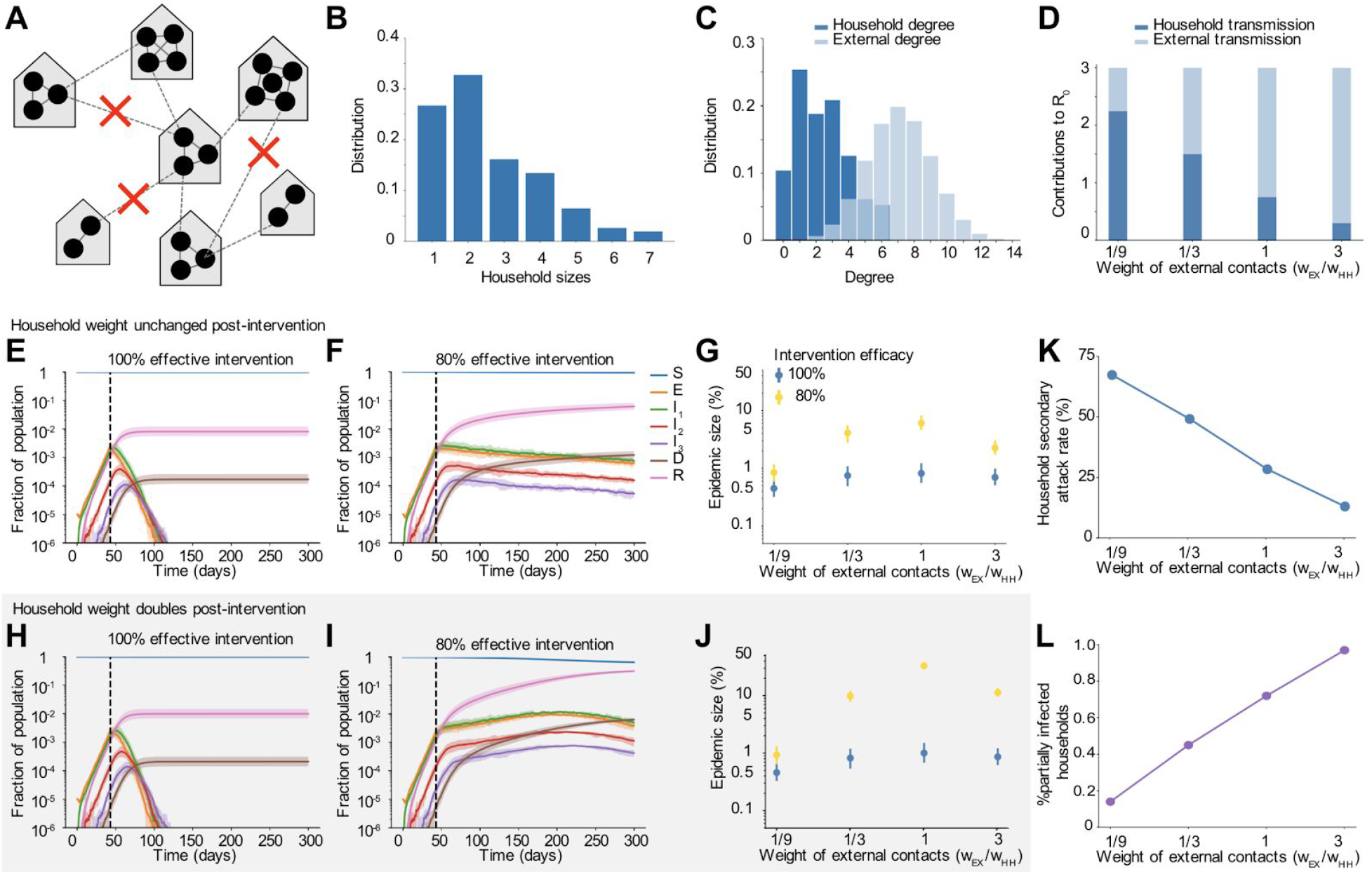
Dynamics pre and post social distancing interventions in network-structured populations with household and external transmission, with neighborhood clustering. A) Multi-layer network of transmission. Individuals have contacts within their households and with others outside the household, which preferentially occur within a local neighborhood (Figure S1). Household and external contacts may have different weights (e.g. different likelihood of transmission), due to for example different levels of physical contact or time spent together per day. Social distancing interventions (red X) remove or decrease the weight of external contacts. B) Distribution of household sizes. C) Distribution of the # of contacts (degree) within the household and outside the household. D) The contribution of household and external spread to the total R_0_ value as a function of the relative weight of external contacts. E)-F) Simulated time course of different clinical stages of infection under an intervention with efficacy of 100% (E) or 80% (F) at reducing external contacts, when household and external contacts have equal weight. Black dotted line shows the time the intervention began. G) The role of the relative importance of household vs external contacts in determining the outcome of the intervention, measured by the size of the epidemic. Epidemic final size is defined as the percent of the population who have recovered by day 300. H-J) Same as above but under the scenario where the weight of household contacts doubles post-intervention (*w*_*HH*_ → 2*w*_*HH*_, due to increased time spent in house). K) The household secondary attack rate, defined as the probability of transmission per susceptible household member when there is a single infected individual in the house, as a function of the relative weight of external contacts. L) The percent of households which are “seeded” with infection at the time the intervention was implemented (i.e. have at least one infected individual). In all scenarios the overall infection prevalence at the time intervention was started was identical.

**Figure S3:**
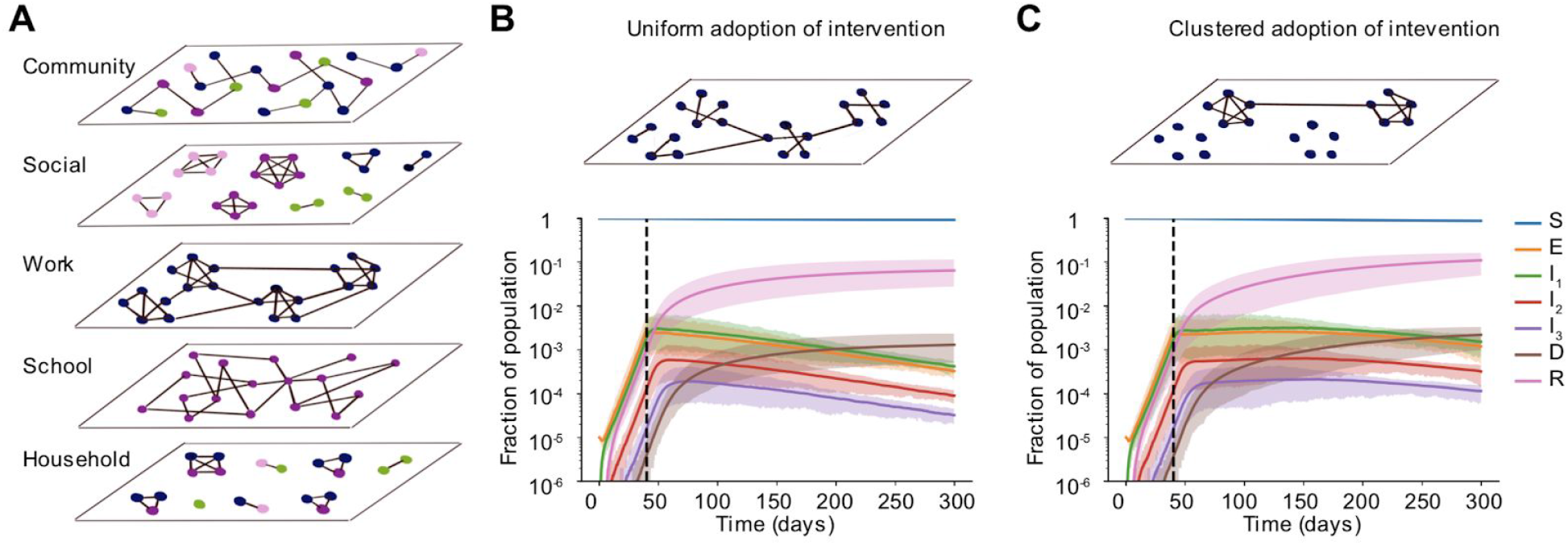
Clustered vs uniform adoption of social distancing measures, with lower efficacy. A) Schematic of the multi-layer network created to more realistically capture non-household contacts and how they are altered by social distancing measures. In each layer, the degree distribution and level of clustering were chosen to match data. The “community” layer represents any other contact not fitting in the other four categories. Colors of nodes represent four broad age groups that determine network membership and structure: preschool-aged (pink), school-aged (purple), working-aged (blue) and elderly (green). B) - C) Simulated time courses of infection in the presence of social distancing intervention with random (B) vs clustered (C) adherence to measures. In both cases, all school connections were deleted post-intervention and 70% of connections were uniformly deleted at random in the social and community layers. In B) 70% of work connections were uniformly deleted whereas, in C) 70% of workplaces were dissolved, leading to clusters of disconnected vs connected individuals in the work layer of the network. The effective intervention efficacy for all layers combined was ∼ 75% in both scenarios. Black dotted line shows the time the intervention began.

**Figure S4:**
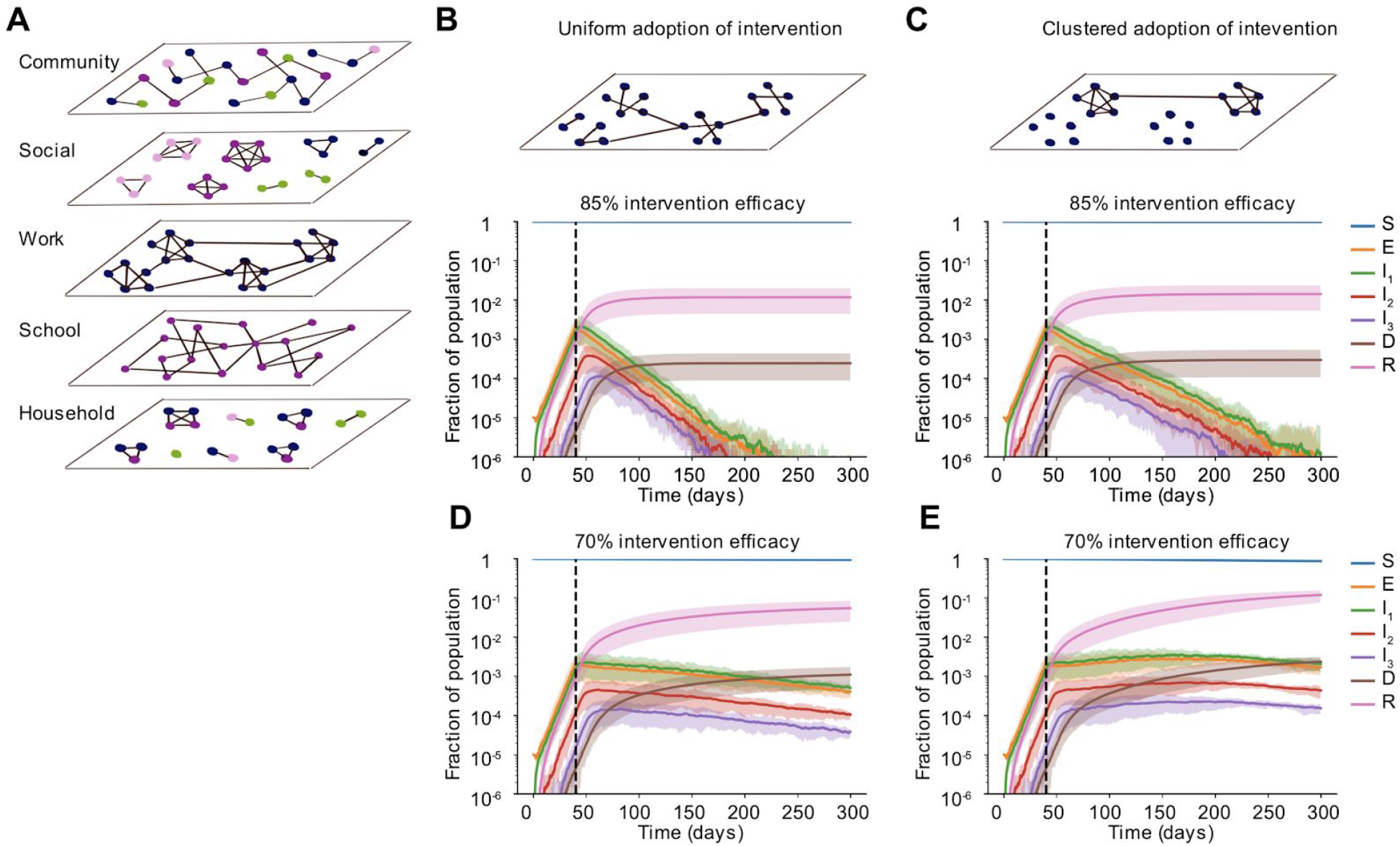
Clustered vs uniform adoption of social distancing measures, with neighborhood clustering. A) Schematic of the multi-layer network created to more realistically capture non-household contacts and how they are altered by social distancing measures. In each layer, the degree distribution and level of clustering were chosen to match data. The “community” layer represents any other contact not fitting in the other four categories. Colors of nodes represent four broad age groups that determine network membership and structure: preschool-aged (pink), school-aged (purple), working-aged (blue) and elderly (green). Community and school connections occurred within local neighborhoods. B) - E) Simulated time courses of infection in the presence of social distancing intervention with random (B,D) vs clustered (C,E) adherence to measures. In both cases, all school connections were deleted post-intervention and 85% (top) or 70% (bottom) of connections were uniformly deleted at random in the social and community layers. In B) 85% (top) or 70% (bottom) of work connections were uniformly deleted whereas, in C) 85% (top) or 70% (bottom) of workplaces were dissolved, leading to clusters of disconnected vs connected individuals in the work layer of the network. The effective intervention efficacy for all layers combined was ∼ 88% (top) or ∼75% (bottom) in both scenarios. Black dotted line shows the time the intervention began.

**Figure S5:**
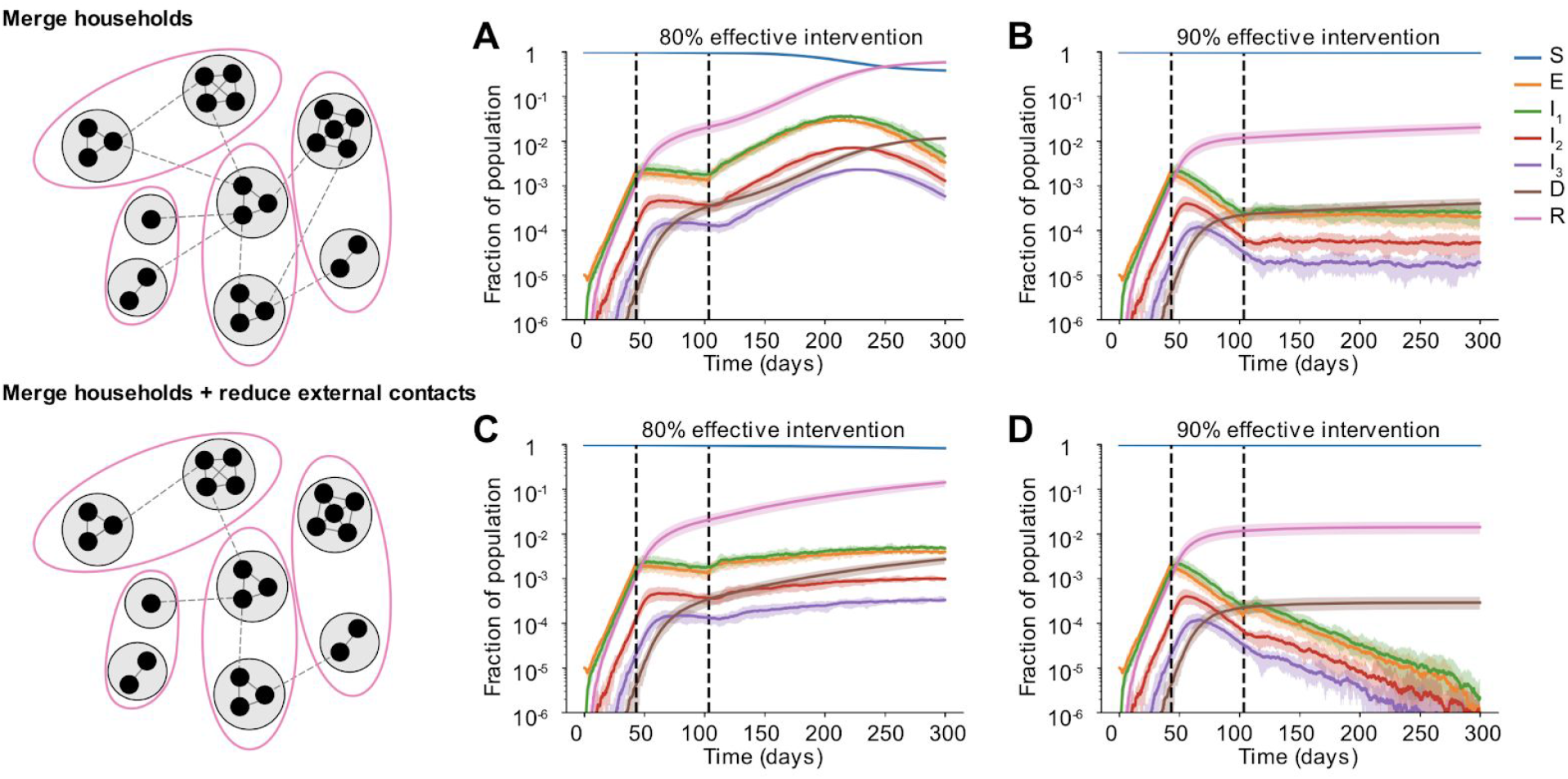
Effect of partially relaxing intervention by forming household bubbles, with neighborhood clustering. Some time after a social distancing intervention was implemented, each household merges with another random household irrespective of their “neighborhood”. In each resulting two-household “bubble”, all individuals are connected to all other individuals. A)-D) Simulated time courses of infection before and after social distancing interventions (with 80% vs 90% intervention efficacy) and after partial-relaxation by household merging. Top row: External contacts of individuals were unchanged after two households were merged, such that overall number of contacts increased. Bottom row: External contacts for individuals were reduced after two households were merged, such that overall number of contacts remained unchanged. In all cases, intervention was started 43 days after the onset of the epidemic (first black dotted line) and was relaxed after two months (60 days, second black dotted line).

**Figure S6:**
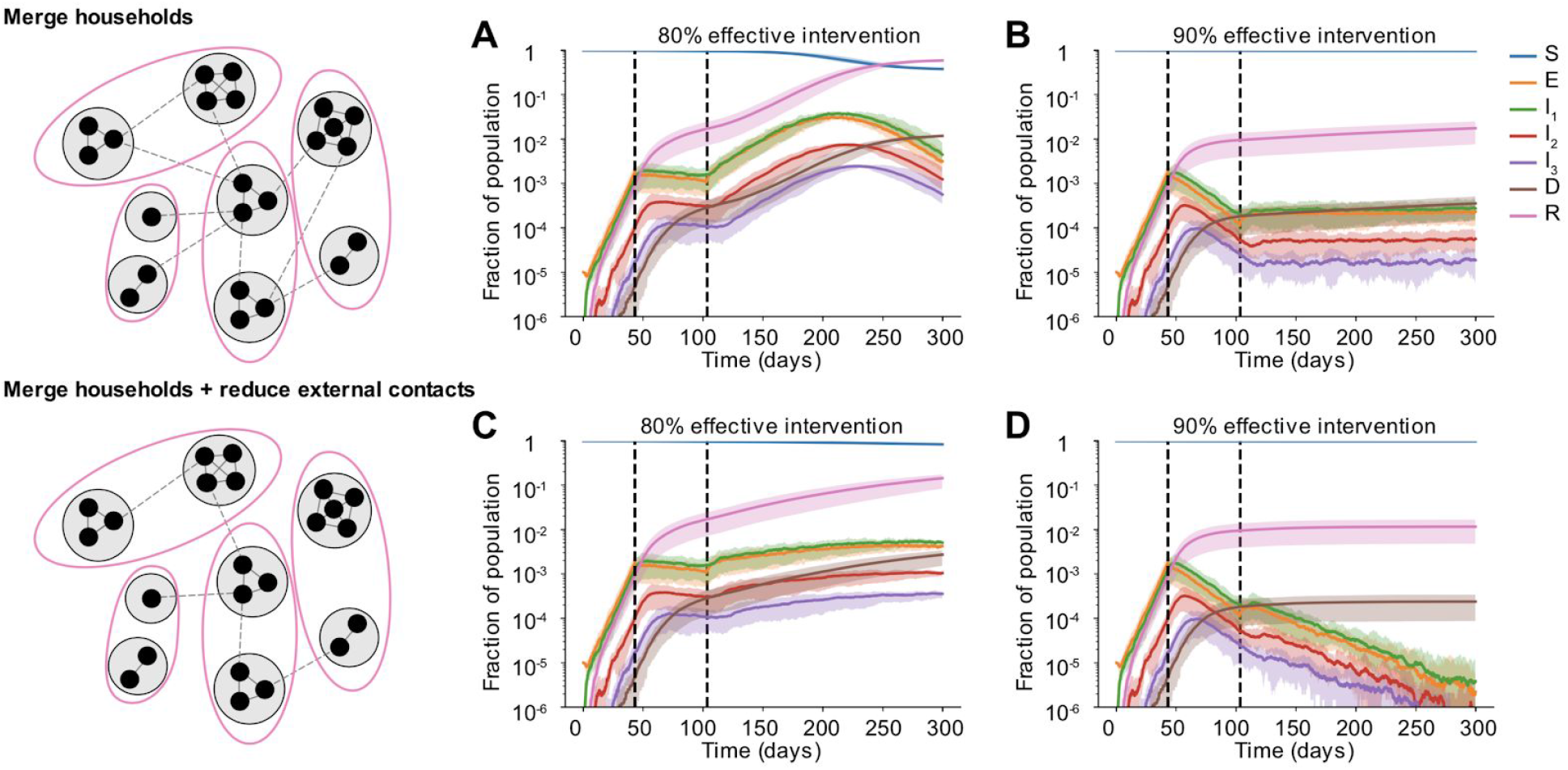
Effect of partially relaxing intervention by forming household bubbles, with neighborhood clustering. Some time after a social distancing intervention was implemented, each household merges with another household from their own “neighborhood”. In each resulting two-household “bubble”, all individuals are connected to all other individuals. A)-D) Simulated time courses of infection before and after social distancing interventions (with 80% vs 90% intervention efficacy) and after partial-relaxation by household merging. Top row: External contacts of individuals were unchanged after two households were merged, such that overall number of contacts increased. Bottom row: External contacts for individuals were reduced after two households were merged, such that overall number of contacts remained unchanged. In all cases, intervention was started 43 days after the onset of the epidemic (first black dotted line) and was relaxed after two months (60 days, second black dotted line).

**Figure S7.**
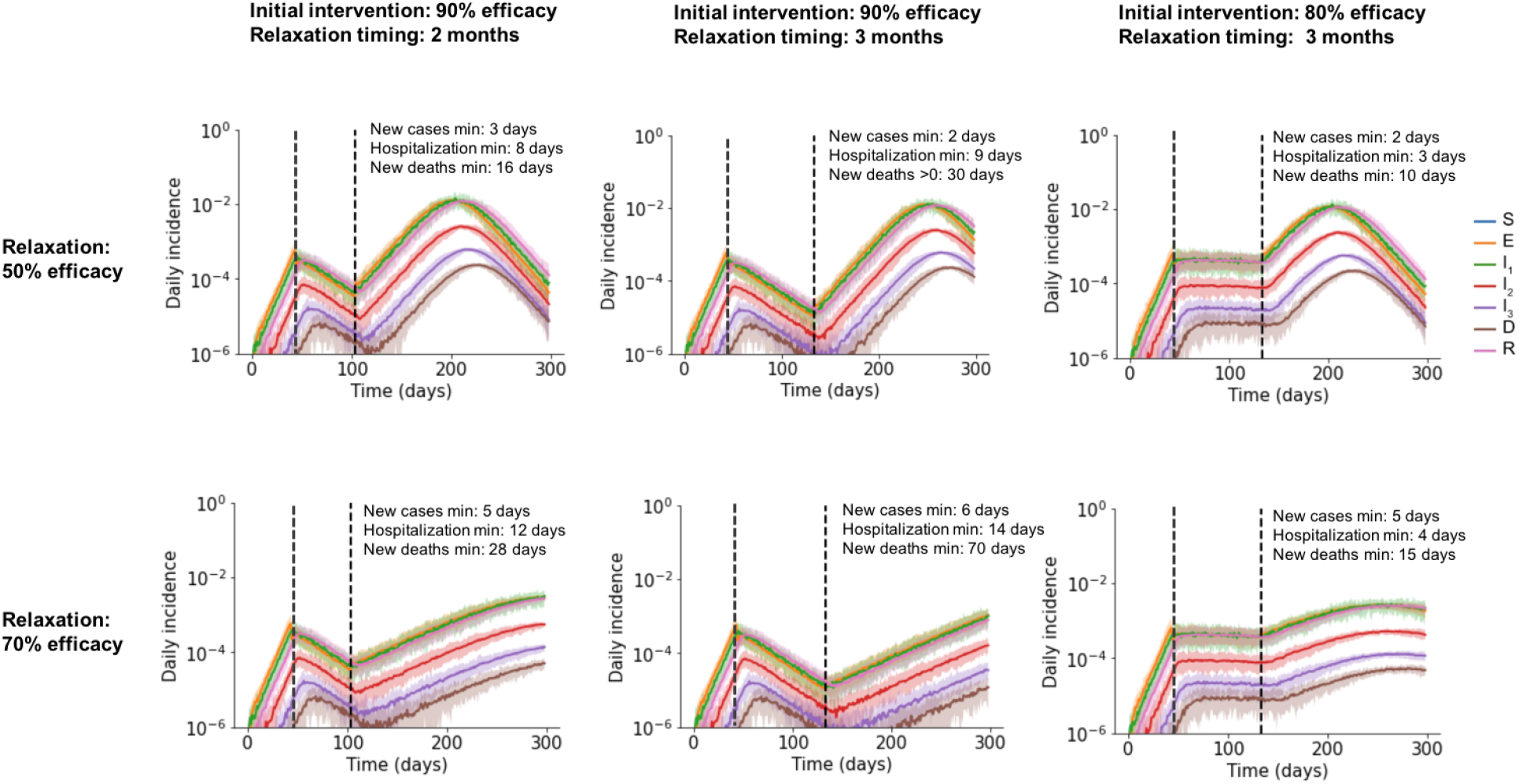
Effects of generalized relaxation of social distancing measures. Simulated time course of different clinical stages of infection under an initial intervention that reduces contacts (first black dashed line) and subsequent relaxation of that intervention (second black dashed line). Plots show daily incidence as a fraction of the total population. Solid line is mean and shaded areas are 5th and 95th percentile. Results are shown for different values of the efficacy of the initial intervention, efficacy during relaxation, and the timing of relaxation. The efficacy of the initial intervention and relaxation are defined as the % reduction in external contacts. In all plots, household and external contacts have equal weight. Plot annotations report the median time post-relaxation until different metrics of disease burden begin to increase - the daily incidence of new cases, current hospitalizations, or daily deaths. If deaths go to zero under an intervention, we instead report median time post-intervention to first death.

**Figure S8.**
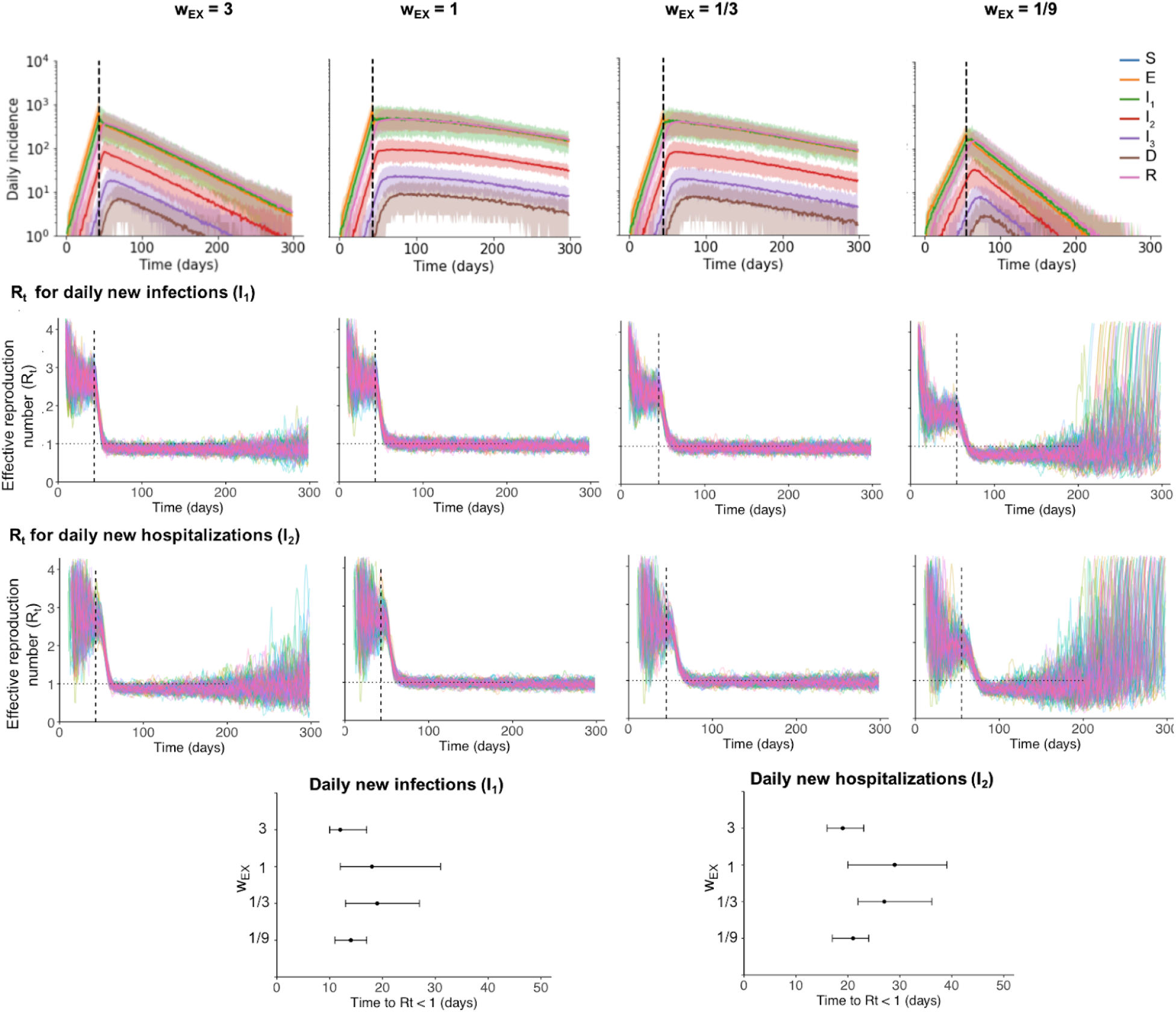
Time-varying effective reproduction number under social distancing. Top row: Simulated time course of different clinical stages of infection under an intervention (first black dashed line) that reduces external contacts by 80%, for different values of the relative weight of external contacts. Plots show daily incidence (for a total population of 1 million). Solid line is mean and shaded areas are 5th and 95th percentile of 100 simulations. Second row: Effective reproduction number (*R*_*t*_) over time calculated using the daily incidence of new infections (I_1_) for each simulation, calculated using the EpiEstim method with a 7-day window. The *R*_*t*_ value plotted for day t is calculated using timepoints before t only. Third row: Same but using the daily incidence of new hospitalizations (I_*2*_). Bottom row: The first timepoint after the intervention that *R*_*t*_ <1. Dots show the median and bars represent 5th and 95th percentiles.

## Notes

### Competing Interest Statement

The authors have declared no competing interest.

### Summary of Updates

Methods section split into Methods and Supplementary Methods. Added an additional analysis that explores the effect of relaxation of social distancing in our model (Suppl. Fig. S7). Added a Supplemental Figure S8, where we calculate Rt from our incidence data using the 'EpiEstim' package.

